# SARSeq, a robust and highly multiplexed NGS assay for parallel detection of SARS-CoV2 and other respiratory infections

**DOI:** 10.1101/2020.10.28.20217778

**Authors:** Ramesh Yelagandula, Aleksandr Bykov, Alexander Vogt, Robert Heinen, Ezgi Özkan, Marcus Martin Strobl, Juliane Christina Baar, Kristina Uzunova, Bence Hajdusits, Darja Kordic, Erna Suljic, Amina Kurtovic-Kozaric, Sebija Izetbegovic, Justine Schaefer, Peter Hufnagl, Alexander Zoufaly, Tamara Seitz, VCDI, Manuela Födinger, Franz Allerberger, Alexander Stark, Luisa Cochella, Ulrich Elling

## Abstract

During a pandemic, mitigation as well as protection of system-critical or vulnerable institutions requires massively parallel, yet cost-effective testing to monitor the spread of agents such as the current SARS-CoV2 virus. Here we present SARSeq, *saliva analysis by RNA sequencing*, as an approach to monitor presence of SARS-CoV2 and other respiratory viruses performed on tens of thousands of samples in parallel. SARSeq is based on next generation sequencing of multiple amplicons generated in parallel in a multiplexed RT-PCR reaction. It relies on a two-dimensional unique dual indexing strategy using four indices in total, for unambiguous and scalable assignment of reads to individual samples. We calibrated this method using dilutions of synthetic RNA and virions to show sensitivity down to a few molecules, and applied it to hundreds of patient samples validating robust performance across various sample types. Double blinded benchmarking to gold-standard quantitative RT-PCR performed in a clinical setting and a human diagnostics laboratory showed robust performance up to a Ct of 36. The false positive rate, likely due to cross contamination during sample pipetting, was estimated at 0.04-0.1%. In addition to SARS-CoV2, SARSeq detects Influenza A and B viruses as well as human rhinovirus and can be easily expanded to include detection of other pathogens. In sum, SARSeq is an ideal platform for differential diagnostic of respiratory diseases at a scale, as is required during a pandemic.

## Introduction

Within just a few months, the newly emerged coronavirus SARS-CoV2 caused the global COVID-19 pandemic^1^. While the world awaits effective vaccines and antiviral therapies, several measures can prevent spread of the virus. Social distancing and more strict “lockdown” strategies are effective in containment but have a major negative impact on human well-being^2,3^. Therefore, the limited and directed application of such measures is desirable. Molecular testing for the presence of the virus by contact tracing and widespread surveillance of asymptomatic individuals, in particular for system relevant institutions and vulnerable person groups, can identify infection clusters and provide the information needed for directed quarantine or other containment measures^4–6^. Such massive testing has shown tremendous impact on containment of the spread of SARS-CoV2 in China, South Korea, Taiwan and Singapore^7–9^.

Several methods have been put forward for assessing infection status, most of which rely on detecting the presence of viral RNA in swab, pharyngeal lavage (gargle), sputum, bronchoalveolar lavage, or saliva samples^10–14^. Tests for the virus itself typically rely on the detection of characteristic fragments of the viral genome or transcripts by reverse transcription (RT) and quantitative polymerase chain reaction (qPCR). Given that PCR reactions can amplify unspecific fragments (incorrect amplicons) despite the use of specific primer pairs, widely used qPCR tests for COVID-19 use fluorescently labeled so-called TaqMan probes that signal the presence and abundance of matching amplicons only. This typically means that one or a few (2-3) amplicons can be detected per reaction, and that specific light cyclers are needed that can perform both PCR and fluorescence measurements. The scalability of such a method is limited by cost and equipment availability – primarily light cyclers.

A more scalable and cost-effective alternative is to couple the same RT-PCR reaction to next generation sequencing (NGS) as a means of high-throughput readout. NGS-based approaches detect amplicons identity by sequencing and computational analyses and therefore are not limited in the number of different amplicons they can detect in parallel: multiple different fragments (viral and cellular controls) can be amplified per reaction, as long as primer pairs are compatible. In addition to detecting multiple fragments in parallel, individual samples can be uniquely labeled with characteristic sequence-identfiers, i.e. indices, to allow for pooled sequencing and subsequent deconvolution. The advantages of detecting multiple pathogen amplicons per sample and processing tens of thousands of samples in parallel mean that NGS-based protocols offer huge cost-saving potential and are thus highly attractive for large-scale testing.

However, while NGS-protocols are conceptually simple and indeed a few different protocols have been developed and partly even FDA-approved^15–20^. Each of these methods have different strengths, yet also suffer from one or several challenges that directly impact sensitivity, specificity at the amplicon and sample level, scalability and/or costs. In this work, we describe SARSeq (*Saliva Analysis by RNA Sequencing*), a robust high-throughput protocol that overcomes these challenges by optimization of the initial sample conditions, a 2-step endpoint RT-PCR, NGS-compatible amplicons with mutually compatible sets of primers, and a barcoding strategy that achieves perfect sample-recall by redundant dual indexing while scaling to tens of thousands of samples by combinatorial indexing along two dimensions. We apply this protocol to samples with synthetic RNAs and various different patient samples and demonstrate that it extends to the simultaneous detection of SARS-CoV2, influenza viruses, and human rhinoviruses (HRV) from the same sample in a single experiment. Overall, our pipeline can be efficiently combined with high-throughput sample collection in 96-well formats, robotics and NGS to detect SARS-CoV2 and other respiratory pathogens in tens of thousands of samples per experiment with a turnaround time of about 1 day (**Fig. 1A**).

**Figure 1.**
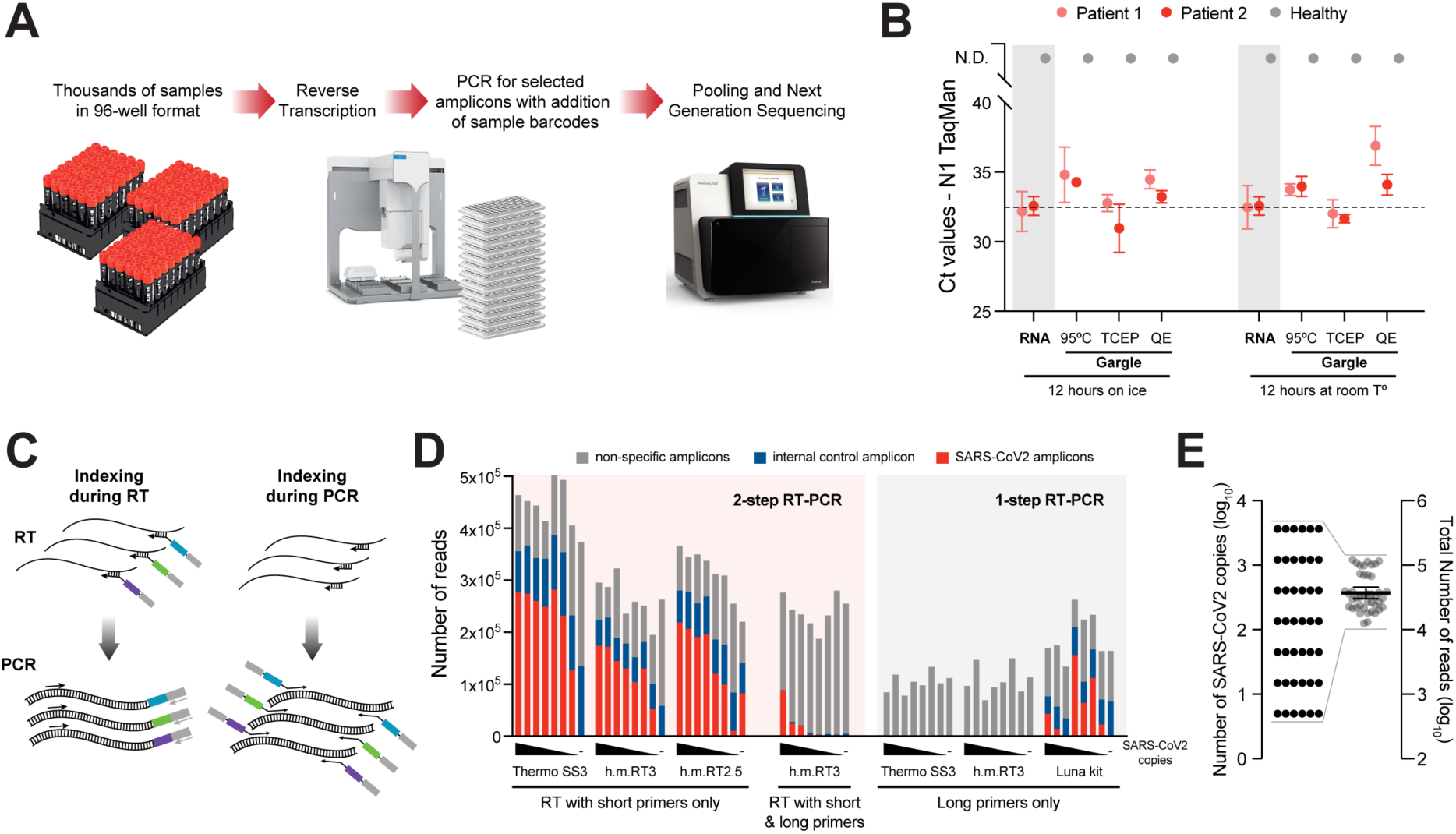
Two-step RT-PCR coupled to NGS allows specific detection of SARS-CoV2 from crude respiratory samples. **A**. Overview scheme to illustrate the envisioned analysis pipeline. **B**. Comparative analysis of SARS-CoV2 detection by TaqMan RT-qPCR on the N1 amplicon, following different RNA extraction treatments. Gargle samples from two SARS-CoV2 positive and one negative person were collected in HBSS. RNA was purified (without concentrating) or crude lysates were produced using either heat inactivation for 30 min^15^, TCEP/EDTA (HUDSON buffer)^22^, or QuickExtract^23^. Treated samples were incubated overnight at room temperature or on ice to assess stability. **C**. Illustration of amplicon indexing either during RT or PCR. Colored boxes symbolize different indices that label individual samples for identification by sequencing (by NGS). Arrows illustrate oligonucleotide hybridization to RNA during RT or DNA in PCR. **D**. Comparison of amplicon levels measured by NGS after indexing either during RT or PCR. Amplicon-specific primers carrying extensions which contain the sample indices (“long primers”) were either included as RT primers (indexing during RT) or used only in the PCR step (indexing during PCR); in the latter case, RT was performed with a mix of random hexamers and two N gene-specific primers (“short primers”) (**Suppl. Fig. 1**). Indexing during RT was performed by either 1-step RT-PCR (with hot start Taq polymerase already present during RT), or 2-step RT-PCR (with hot start Taq polymerase added after RT). Black wedges symbolize a dilution series of synthetic SARS-CoV2 RNA from 3645, 1215, 405, 135, 45, 15, 5, to 0 molecules. Thermo SS3: Superscript III enzyme provided by Thermofisher; h.m. RT3: homemade Superscript III-like enzyme; h.m. RT2.5: homemade variant of Superscript II-like enzyme; **E**. Total NGS read numbers per individual sample, for a set of 42 samples containing from 5 to 3645 SARS-CoV2 RNA molecules; note the range compression from four to one order of magnitude, which enables equal representation of all samples across the sequencing space and therefore high sensitivity and scalability.

## Method Development and Results

### Two-step RT-PCR allows specific detection of SARS-CoV2 from crude respiratory samples by NGS

The first step towards establishing a high-throughput SARS-CoV2 test was to find a sample preparation method that would bypass the costly and time-consuming step of RNA purification from patient samples, while being compatible with RT-PCR. A number of sample types have been effectively used to detect SARS-CoV2, including swabs collected in viral transport medium (VTM) or other buffers, saliva and gargle with Hanks’ balanced salt solution (HBSS) or saline solutions^10–12,14^. Gargle samples enable similar sensitivity to swabs collected by medical staff^12^, and are preferred to pure saliva as samples become more uniform in viscosity and are thus easier to pipette, a prerequisite for automation (**Fig. 1A**). Such samples, however, pose the challenge that exposure of viral or cellular RNA for RT must occur under strict inhibition of the high load of RNAses present in saliva^21^. A number of methods have been reported to expose and simultaneously stabilize RNA in these samples, including heat inactivating at 95°C^15^, treating with proteinase K^14^, and mixing with TCEP/EDTA^22^ or with QuickExtract solution^23^. To compare these methods, we obtained gargle samples (in HBSS) from one negative and two SARS-CoV2 positive individuals, and either purified RNA or treated the gargle according to the different protocols. We then assessed RNA exposure and stability by performing TaqMan RT-qPCR of a virus-specific amplicon (N1) (**Fig. 1B**). All the methods generated stable RNA while maintaining similar sensitivity to purified RNA under our reaction conditions. We also tested QuickExtract and TCEP/EDTA on swab samples in VTM, in experiments that are described below. In these tests, QuickExtract showed the least precipitation upon heating to 95°C and was thus used for most experiments unless otherwise stated.

The next aspect we evaluated was when to add the DNA indices that distinguish individual samples. These can in principle be incorporated during the RT^15,18,19^ as well as during the PCR^24,25^, as extensions of the primers used to reverse transcribe or amplify the desired amplicons, respectively **(Fig. 1C)**. However, we found that having primers with the required extensions during the RT step resulted in a large fraction of non-specific PCR products, presumably because the low temperature of the RT reaction allows substantial non-specific priming (**Fig. 1D**). Given the large but limited sequence space on NGS flow cells, such lack of specificity means that many more reads would be needed per sample, limiting upscaling to large sample numbers. We therefore chose a 2-step reaction in which priming in the RT step is performed with random hexamers plus two gene-specific 12-mers that increase sensitivity for SARS-CoV2 (**Suppl. Fig. 1**), while integration of the sample indices occurs during the PCR.

One of the hurdles towards establishing a pooled NGS-based assay for samples from virus infected individuals derives from the fact that viral loads can differ by many orders of magnitude such that high-titer samples would dominate an NGS run. TaqMan RT-qPCR reports differences in Ct values of 20-25 cycles, which translate into 2^25^=33.5 million-fold differences in viral titers. Therefore, if samples with low virus titer are to be robustly identified as positive, e.g., with >100 virus-derived amplicon reads, the samples with high virus titers would require 3.3 × 10^9^ reads, which is prohibitive. For that reason, the dynamic range needs to be compressed to “dampen” the signals from highly positive samples while providing sufficient sensitivity to detect samples with lower titers. Of note, sample indexing during RT as discussed in Fig 1C with subsequent PCR upon pooling would also maintain these quantitative differences, which is not desired for this application^19^. To achieve this compression, we ran the first PCR reaction on individual samples for 45 cycles until they reached saturation. This generated similar numbers of amplicons per well independent of initial viral titer (**Fig. 1D,E; Suppl. Fig. 1C**). In summary, using crude respiratory specimens as input, a 2-step end-point RT-PCR generates high-specificity and uniform representation of correct amplicons across samples and enables pooling of many samples for analysis by NGS.

### A control primer pair targeting 18S rRNA provides better specificity than the widely used RPP30 primers

In addition to the very large dynamic range of viral titers between patients, non-specific PCR amplicons can impair the detection of viral amplicons by NGS, because the number of NGS reads is inherently limited (and directly proportional to the total costs). For example, the parallel analysis of approximately 40,000 (96 × 384) samples means that each sample can receive a total of ∼500 reads on a MiSeq, ∼2,000 reads on a HiSeq, and ∼10,000 reads on a NextSeq platform. If a substantial fraction of these reads were spent on sequencing non-specific amplicons, assay sensitivity would be severely impacted. It is thus pivotal to select amplicons and primer pairs that i) show high sensitivity, ii) generate amplicons of comparable short size, and iii) generate few non-specific amplicons alone or in combination with any other primer present in the same reaction, which is of particular importance when using primers with long extensions (here: up to 42 nucleotides as PCR primers contain sample-identifying index sequences, staggers of random nucleotides, and primer-binding sites for a 2nd PCR as discussed below).

We thus tested several published SARS-CoV2 specific primer pairs^26–29^ after adding our index-containing extensions. We settled on the N gene-specific primer pairs N1 and N3 proposed by the Center for Disease Control (CDC) as they produce an ideal amplicon length of ∼70 bp and performed best in SYBR-Green qPCR (which does not control for amplicon identity) as well as in initial sequencing runs (**Suppl. Fig. 2**). We then tested the N1 amplicon together with the widely used internal control primer pair targeting *RPP30* (coding for RNAse P). While the N1 primers showed up to 50% of correct amplicons, in correlation with the amount of synthetically spiked in template, the fraction of specific amplicons for *RPP30* was only 0.06-1.5% (**Fig. 2A**). When analyzing all sequenced amplicons across all samples shown in 2A, we detected various short sequences that together made up >99% of all NGS reads. The vast majority was generated by the *RPP30*-specific primers (**Fig. 2B**), suggesting that these primers are not compatible with multiplexed PCR and NGS. We therefore set out to establish a new control primer pair that would produce fewer non-specific amplicons. We tested several primer pairs on gargle samples obtained from 16 individuals, yet only a single primer pair, specific for 18S ribosomal RNA, was detected in all samples and showed a strong dependency on the presence of reverse transcriptase (**Fig. 2C**). Ribosomal amplicons were detected at Ct values of 15-45, a range comparable to that of viral amplicons in infected individuals. We therefore tested ribosomal RNA as a host control in our SARSeq panel. Indeed, the ribosomal amplicon performed well and neither generated abundant unspecific amplicons alone, nor with N1 or N3 primer pairs (**Fig. 2D**).

**Figure 2.**
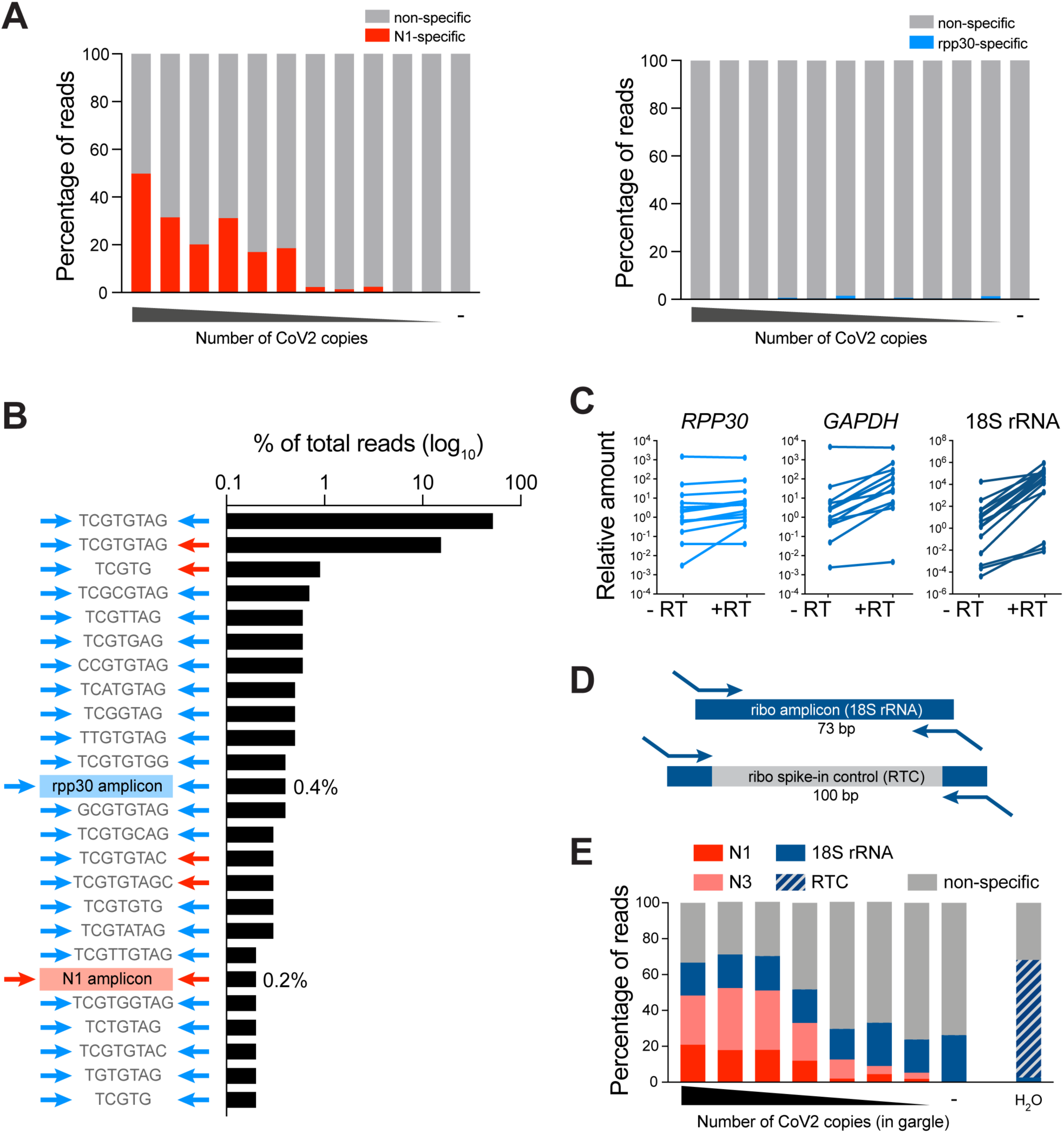
A control primer pair targeting 18S rRNA improves assay performance compared to the widely used *RPP30* primers. **A**. NGS read counts obtained with the SARS-CoV2 N gene-specific primer pair N1 as well as the human RNAse P (*RPP30*)-specific primer pair designed by the CDC^28^. RT-PCR was performed on inactivated gargle-QE samples spiked with a range of 5120 to 5 molecules of synthetic template in a two-fold dilution series, as well as 0 molecules. Non-specific amplicons are defined as amplicons generated by the respective matched primer pairs but with incorrect sequence in between. **B**. Analysis of all amplicons generated in the pool of conditions shown in A. Non-specific amplicons typically incorporated a short stretch of sequence complementary to some primer sequences and were almost exclusively generated by at least one *RPP30*-specific primer. Specific amplicons add up to <1% of reads in this condition. **C**. SYBR Green qPCR analysis of *RPP30* primers and two alternative internal control primer pairs in the presence and absence of reverse transcriptase on RNA purified from gargle of 16 individuals. Ct values are transformed to relative differences. **D**. Scheme of the RT control (RTC) spike-in; an amplicon with identical primer binding sites to the ribosomal internal control yet different and longer intermediate sequence was synthesized, cloned, and T7 transcribed. The RTC was added during the RT at 1000 molecules/reaction. The ratio of ribosomal to RTC reads serves as a sample quality measurement. Reactions without reads for the ribosome or the RTC amplicons indicate an inhibited/failed RT-PCR reaction. **E**. Read distribution from an NGS experiment similar to A, but with the ribosome amplicon as internal control, and addition of the RTC spike-in. The number of non-specific reads is dramatically reduced, which impacts sensitivity and scalability.

Since the 18S amplicon - like the *RPP30* amplicon - does not span an intron, it cannot discriminate against genomic DNA templates abundant in respiratory samples. We thus designed an additional internal RNA control to assess successful reverse transcription in all samples (reverse transcription control or RTC); we produced an in vitro-transcribed RNA with identical primer binding sites as the ribosomal amplicon, yet an unrelated sequence in between **(Fig. 2E)**. The RTC was spiked into the reverse transcription mix at a concentration of 1000 molecules/reaction **(Fig. 2D)**. Upon further optimization of RT- and PCR buffers **(see Methods)**, SARSeq reached high amplicon specificity with >30% amplicons corresponding to expected amplification products **(Fig. 2D, see also Fig. 1D)**. Moreover, the ratio between ribosomal amplicon and RTC reads provided a good assessment of sample quality: we observed that in the presence of good-quality gargle, the ratio is high and the RTC is lowly detected, but if the sample is low in nucleic acids the RTC takes over and the ratio is low. We also encountered clinical samples that likely contained RT and/or PCR inhibitors in which neither amplicon was efficiently detected (not shown). The high specificity of amplicons achieved, and the even representation of reads across samples set the stage to develop a high-throughput indexing strategy that allows analysis of tens of thousands of samples in parallel.

### Two-dimensional redundant dual indexing allows scaling to population-level testing

To exploit the high-throughput nature of NGS, we would need a sample barcoding strategy that allows multiplexing of tens of thousands of samples in a single sequencing run while retaining strict sample specificity, i.e., suppressing misassignment of reads to incorrect samples, which can lead to false positive diagnoses. Several strategies for sample indexing are possible. First, samples can be individually indexed by a sample-specific short DNA sequence (typically called *index* or *barcode*) in the RT primer or one of the two PCR primers (**Fig. 3A**). In such a setup, sample-specific primers incorporate sample-indices into all amplicons from each sample, and these are then sequenced as part of the respective amplicon; e.g., Salis and colleagues designed 19,000 RT indices^19^. This strategy, however, does not scale well as it requires distinct primers for each amplicon and sample (linear/additive scaling). More importantly, it cannot retain perfect sample identity due to template-switching PCR artifacts^30,31^ and index-hopping on flow cells^32,33^, which can lead to incorrect associations between amplicon- and index sequences. This problem is of particular relevance when high-titer samples are analyzed next to samples from healthy individuals as we demonstrated by spiking synthetic SARS-CoV2 template into two wells of a 96 well plate (wells B8 and F2) in which all other wells are negative (**Fig. 3B**). The scalability limitation can be overcome with a combinatorial indexing strategy, such as a column index on the forward primer and a row index on the reverse primer^34^ (combinatorial/multiplicative scaling; **Fig. 3C**). However, such a strategy suffers from the same inability to retain perfect sample identity, which in this case leads to a characteristic cross-shaped pattern along the rows and columns of the positive samples due to the misassignment of the row or column indices (**Fig. 3D**).

**Figure 3.**
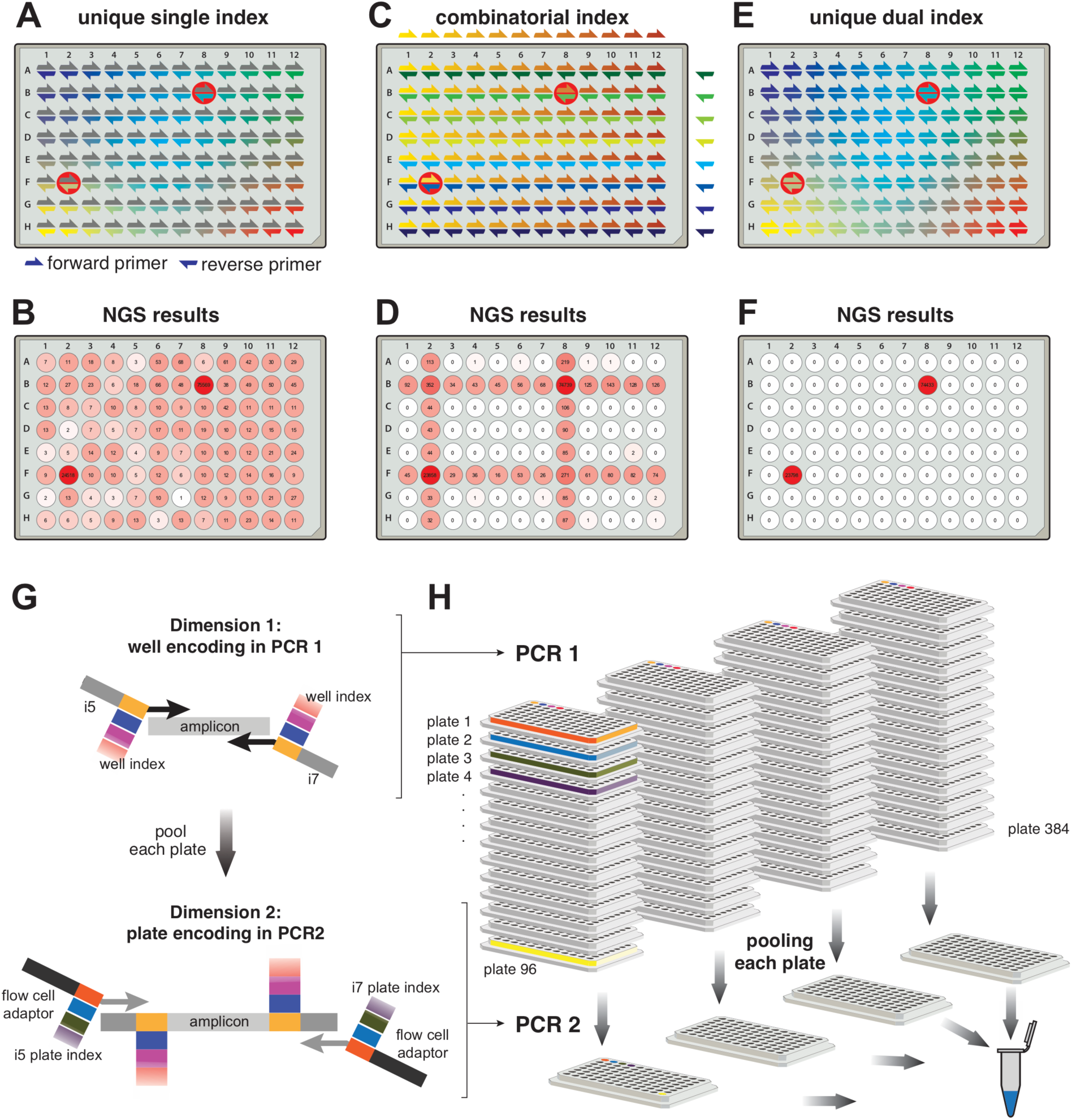
Two-dimensional redundant dual indexing allows scaling to population-level testing. **A**. Scheme depicting 96 well-specific indices. These can be incorporated by forward or reverse primers. For the latter they can be incorporated during RT or PCR. Red circles highlight the positions into which synthetic SARS-CoV2 RNA was added to test specificity of the indexing strategy. **B**. N3 amplicon reads obtained by NGS and mapped to each well based on indices incorporated by reverse primers during PCR. Forward primer indices were disregarded in this analysis. Note the frequent mis-assignment to incorrect positions. **C**. Scheme depicting combinatorial indexing. Each well is identified as a unique combination of a forward and a reverse index. **D**. N3 NGS reads mapped to wells based on combinatorial indexing as in C. To simulate combinatorial indexing the identical dataset as in B and F was used and primers were treated in pools pointing to columns or rows. **E**. Unique dual indexing is a redundant indexing method that encodes each well both by a unique forward and a unique reverse index. Thus, illegitimate recombination products between an amplicon and its associated indices can be bioinformatically rejected. **F**. NGS result for the same dataset as in B and D analyzed with unique dual indices successfully filtered away all misassigned reads. **G**. Two rounds of unique dual indexing (in two subsequent PCR reactions) can be used to index first wells and then plates, effectively achieving combinatorial (multiplicative) scaling. Colored boxes represent indices. **H**. Illustration of the PCR workflow. RT and PCR 1 are performed on all samples individually, adding well-specific indices. Subsequently each plate is pooled to one well position of a new plate, reactions are treated with Exostar to remove excess primers and a second PCR is done, adding plate-specific indices and the Illumina flow cell adaptors. Our currently used and validated index set allows pooling of up to 36,864 samples (96 in PCR1 x 384 in PCR2). Finally, PCR 2 amplicons are pooled, gel purified and sequenced on any Illumina platform.

We developed an indexing scheme for SARSeq that achieves perfect sample specificity and combinatorial scalability. Specificity regarding sample identity was achieved by two indices that both point to the same sample/well (**Fig. 3E, F**), a strategy termed *redundant dual indexing* or *unique dual indexing*^34,35^. These two indices are introduced through forward and reverse primers and redundantly encode each sample with distinct indices at each end of the amplicon, thereby eliminating illegitimate index combinations. Such an approach requires two indices (=unique primers) per sample and therefore does not scale well when a single PCR is used (one dimension). We therefore use a two-dimensional indexing strategy, which we realized by two subsequent PCR steps: after the first PCR performed with unique dual indexing, we pool all samples within one plate into one well of a second plate and perform a second PCR that again uses unique dual indexing. This strategy of *two-dimensional redundant dual indexing* allows combinatorial indexing between dimension 1 and 2, and thus multiplicative scaling, while retaining perfect sample identity (**Fig. 3G, H**). It requires an only modestly higher number of indexing primers for very many samples and allows the encoding of 96×96 or 96×384 samples with 2×96 amplicon-specific primers (2 per amplicon; 1st dimension) plus 2×96 or 2×384 global primers (irrespective of amplicon; 2nd dimension), respectively.

In practice, we extended the amplicon-specific primers for the 1st PCR (1st dimension) at their 3’ ends to include a sample-specific index and i5/i7 sequences as primer-binding sites for the 2nd PCR. To ensure sufficiently complex sequences of the NGS forward reads for cluster identification and stable sequencing, we staggered the sample-index and the amplicon-specific sequence by a random offset of 1-4 base pairs **(see Fig. 3G and Suppl. Table 1)**. We tested a total of 110 primer pairs per amplicon (N1, N3 and 18S rRNA) to establish a set of 96 primer pairs that show good amplification behaviour for all amplicons **(Suppl. Fig. 2)**. For the final set of primers, all amplicons within one well obtain identical offsets and indices to prevent recombination between amplicons and indices during PCR and to simplify bioinformatic analysis. Pre-prepared primer-plates and robotic pipetting pipelines allow us to process thousands of samples in parallel.

After the indexing of individual samples (=wells of a 96-well plate; 1st dimension), all samples of one plate were pooled to one position of a new 96-well plate, and in a second PCR, a plate-specific index was added (2nd dimension). We implemented three measures to ensure that sample identity was perfectly retained between the 1st and 2nd PCR. First, to ensure that primers from the 1st PCR were used up in PCR 1 and thus not present during the 2nd PCR, we included an RNA template with N1 and N3 primer binding sites similar to the RTC and the normalization-spike-in used in the Swab-Seq pipeline^15^ (**Fig. 4A**). Second, we treated the pools of PCR 1 with DNA exonuclease to enzymatically degrade all single-stranded DNA and thus all remaining primers especially from SARS-CoV2 negative wells. Third, we kept the cycle number for PCR 2 at a minimum to avoid amplicon recombination during PCR and used a PCR protocol that prevents premature termination of an extension step^36^. Indeed, all three measures synergistically contributed to the robustness of read assignment **(Suppl. Fig. 3A)**. In each dimension we used dual and redundant indices with a Hamming distance of at least three mismatches. The primers used for PCR 2 (2nd dimension) are commercially-available Nextera primer sets available as 384 unique pairs, which are frequently used in many NGS sequencing facilities for multiplexing. This pipeline also suppressed read misassignment across plates, such that the highly-positive positions of Fig. 3A, C, E were not detected on other plates run in the same experiment **(Suppl. Fig. 3B)**.

**Figure 4.**
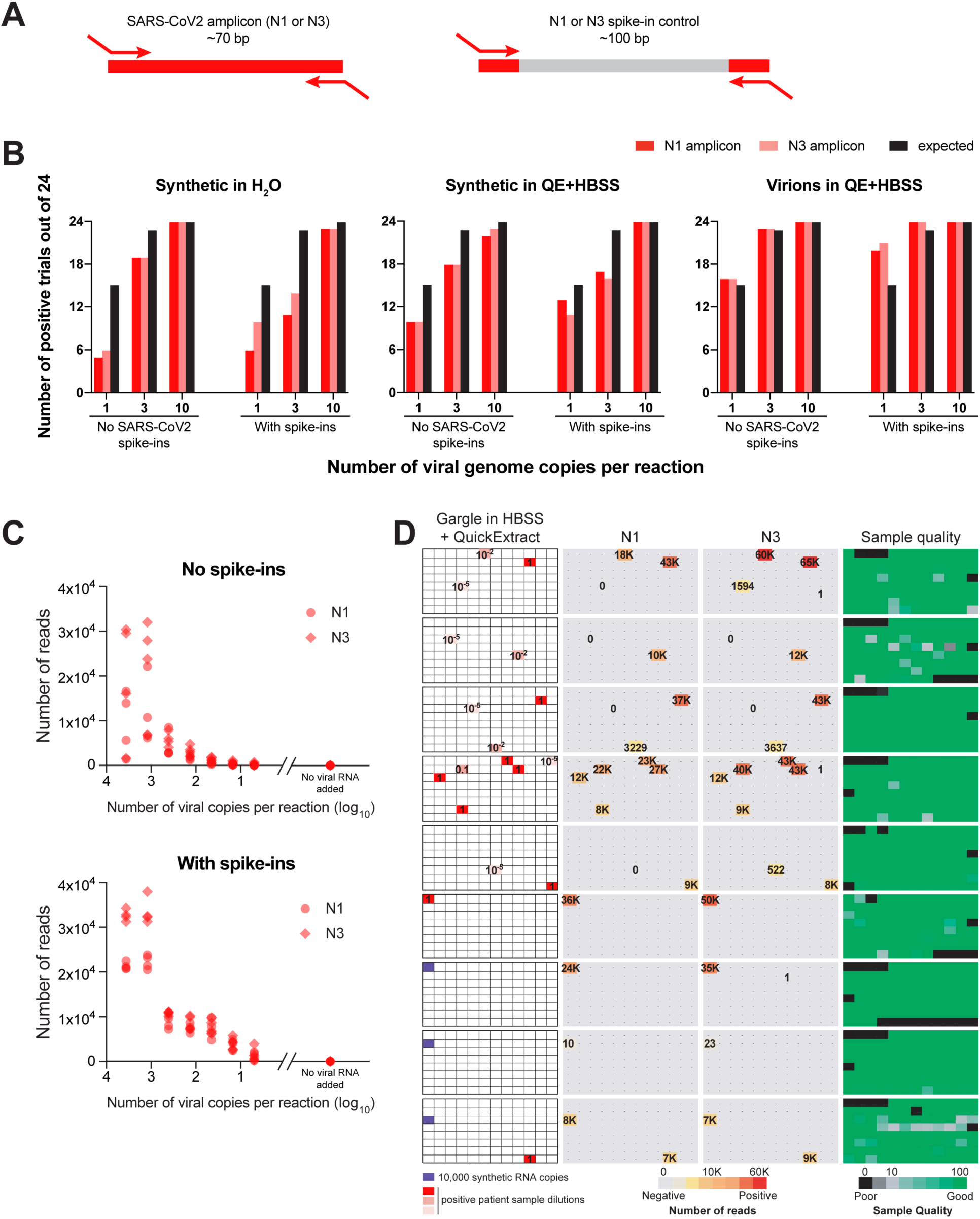
SARSeq is specific and sensitive when tested on a large set of gargle samples. **A**. Schematic illustration of RNA spike-ins to scavenge N1 and N3 primers during PCR 1 in SARS-CoV2 negative samples. Analogous to the RTC, primer binding sites are identical to N1/3 amplicons, but intermediate sequence is distinct and longer so as not to compete with virus-derived amplicons. **B**. Frequency of detection of SARS-CoV2 at minimal template concentration. Number of detected cases out of 24 trials is depicted. Note that the expected frequency of detection is only 63% and 95% for one and three molecules respectively, assuming a Poisson distribution of molecules in wells. **C**. Read counts of N1 and N3 amplicons in a synthetic SARS-CoV2 RNA dilution series performed in four replicates. Note the reduced variability in the presence of N1/3 spike-ins. **D**. SARSeq performance on a test pool of 864 gargle samples collected in HBSS from healthy participants of a routine SARS-CoV2 testing pipeline at our institutes. These were spiked with synthetic SARS-CoV2 RNA or a dilution series generated from a positive patient sample(Ct=30). All negative samples produced 0-1 N1/3 reads, while positive samples produced thousands. The only missed samples were 10^−5^-fold dilutions of the patient sample, which presumably did not contain SARS-CoV2 RNA. Sample quality is assessed by the ratio or ribosomal reads to RTC spike-in.

Our experimental design provides scalable and robust indexing for SARS-CoV2 amplicons by using PCR to incorporate two sets of redundant indices, and bioinformatics to only allow legitimate combinations of those four indices. The ability to encode 96 well indices (1st dimension) and 384 plate indices (2nd dimension) means this can be used to prepare 36,864 individual samples simultaneously. To illustrate scalability of the approach, a 4-fold increase in one dimension for example by using 384 well indices in the 1st PCR, would enable multiplexing of >145,000 samples. This degree of multiplexing renders the sequencing price per sample negligible and thus enables frequent population-wide testing because sequencing capacity is also not bottleneck with current NGS platforms. In summary, redundant dual indexing ensures sample identity specificity and two-dimensional indexing allows scalability while preventing any spill of reads from positive to negative samples even across multiple orders of magnitude in signal intensity.

### SARSeq is specific and sensitive when tested on a large set of gargle samples

To test sensitivity, specificity, and scalability of SARSeq we set out to run large sample cohorts in which we diluted synthetic RNAs or a high-titer patient sample into SARS-CoV2-negative gargle samples from hundreds of different people. In addition, we also used this setup to test the effect of spike-ins with identical primer binding sites to the N1/N3 amplicons but different sequences in between, as introduced previously^15^ **(Fig. 4A)**. We processed multiple 96-well sample plates in parallel using a robotic pipetting platform.

We first assessed the sensitivity of SARSeq in this setup by diluting viral templates to 1, 3, or 10 copies per reaction (0.2-2 copies/µl in the 5 µl sample input). To account for the contribution of QuickExtract as well as to test RNA exposure from viral particles, we diluted synthetic RNA in H2O as well as QuickExtract:HBSS but a l s o v i r i o n s p a c k a g e d i n c e l l c u l t u r e, i n QuickExtract:HBSS. We measured each dilution in 24 replicates using SARSeq as described above. Using H_2_O for dilution we detected the 1 copy solution of SARS-CoV2 RNA in 5 and 6 of 24 tested cases for N1 and N3, respectively. At such dilution, assuming a Poisson distribution, 63% of wells are expected to contain one or more viral copies, pointing towards a detection efficiency of 30-40% per molecule. QuickExtract increased that efficiency to 70% while detection straight from viral particles was at 1.1 per molecule **(Fig. 4B and Suppl. Fig. 4)**. It is thought that infectious COVID-19 patients show viral titers of >10^3^/ µl; our detection limit is thus at least 100 times more sensitive than required for mitigation strategies for the SARS-CoV2 pandemic.

We had designed spike-ins containing the N1 and N3 priming sites to ensure that these primers are used up even in the absence of viral templates (**Fig. 4A**). We wondered if possible primer competition would thus decrease sensitivity of the assay. The presence of 100 copies of each the N1 and N3 spike-ins did not decrease sensitivity but even showed slightly better detection efficiencies, presumably also because the spike-ins were longer than the viral amplicon (**Fig. 4B**).

Due to the endpoint PCR we perform, SARSeq intentionally only returns semi-quantitative results, yet as such spike-ins have been used to improve the quantitative ability of other NGS approaches that rely on endpoint PCR (the ratio between viral amplicon and spike-in reads reflects the ratio of these two templates in the starting reaction^15^), we tested the effect of spike-ins on the quantitative behaviour of SARSeq (**Fig. 4C**). At the level of 100 copies per reaction the number of reads we obtained corresponding to the spike-ins was too low to serve as a reliable “denominator”. Nevertheless, when testing a dilution series of synthetic SARS-CoV2 RNA in quadruplicates, we noticed improved reproducibility in the presence of these spike-ins and therefore included them in the final setup. Also, our assay retains a degree of semi-quantitativeness over three orders of magnitude (**Fig. 4C**).

To challenge SARSeq with hundreds of real samples omitting RNA purification and thus prepare for a clinical performance study, we generated sample plates from pharyngeal lavage (gargle) collected in HBSS from healthy participants of routine SARS-CoV2 testing at our institutes. Such diverse, crude samples may contain reagents inhibitory to the RT or PCR step. All gargle samples were previously tested negative through qPCR but we added synthetic SARS-CoV2 RNA and a dilution series of a positive gargle sample with Ct=30 in a TaqMan qPCR assay, into several marked positions (**Fig. 4D**). Subsequent to PCR 1 we upscaled the experiment by creating 6 replica each sample plate and thus brought it to 180 virtual plates that were processed in parallel in PCR 2 and sequenced on one NextSeq high output lane. We therefore measured a total of 2,880 real and 17,280 replicated samples in this batch. We detected all positive samples, with the exception of a 10^−5^ dilution of a positive gargle sample with a Ct value of 30, suggesting very high sensititivty. Notably, the agreement between N1 and N3 amplicon-based results was 100% **(Fig. 4D)**.

Another critical parameter when testing large numbers of patients is the false positive rate. We were therefore pleased to see that our indexing strategy and pipeline delivered typically zero and very rarely 1 read indicative of SARS-CoV2 for gargle samples previously tested negative by qPCR as well as for all H_2_O controls, compared to hundreds or thousands of reads for positive samples. In total we performed four runs using gargle samples from our in-house testing pipeline, adding up to 4952 negative samples and 728 positive samples created by adding synthetic SARS-CoV2 RNA or dilutions of a positive patient sample. We observed 2 unexpected N1-positive samples and 5 unexpected N3-positive samples, estimating a false positive rate for our pipeline of 0.04-0.1%. This binary result showcased an unambiguous assessment of infection status by SARSeq. Due to the absence of false positive reads we also did not need to further use the N1 and N3 spike-in “denominator” amplicons to set a threshold ratio for calling positive results^15^. In summary, SARSeq enables the semiquantitative assessment of synthetic SARS-CoV2 RNA in various buffers and in gargle samples and allows the detection of SARS-CoV2 RNA with high sensitivity and specificity.

### SARSeq robustly detects SARS-CoV2 in patient samples from a clinical setting

To test if SARSeq robustly detects real SARS-CoV2 virus in patient samples collected in clinical diagnostic settings, we measured a set of 564 swab samples from independent patients collected at the Clinical Center of the University of Sarajevo (Sarajevo, Bosnia and Herzegovina). These samples were obtained in VTM and further mixed with QuickExtract, in duplicates. While we did not detect N1 or N3 amplicons in H_2_O or - RT conditions (**Fig. 5A, B**) we frequently obtained SARS-CoV2 reads across all plates with good correspondence between amplicons and replicates (**Fig. 5C, D**). To assay correspondence to the standard test, we also measured the samples in a TaqMan qPCR assay in duplicates (also without prior RNA purification). As expected, we observed a robust correlation of both qPCR replicates until ∼Ct=36 **(Fig. 5E, red dots)** and stochastic behaviour beyond that detection limit **(Fig. 5E, orange dots)** with either one or two replicates scoring positive. No SARS-CoV2 was detected in an additional 354 samples. We analyzed the overlap between detection by qPCR and NGS and found that 157 (96.3%) of samples that scored with a Ct value of <36 in at least one qPCR scored positive in all four assays, while six samples scored positive in one or two assays only **(Fig. 5F)**. We hypothesized that the stochastic behaviour at the detection border is due to the presence or absence of a single reverse transcribed viral genome. If that was true, detecting that genome by both amplicons in a single replicate should be more likely than detecting one of the two amplicons in the two independent replicates. In contrast, different sensitivies towards the N1 and N3 amplicons would result in the opposite outcome. Indeed, of the samples detected with 1-3 of these assays, 48 and 43 patient samples showed detection of SARS-CoV2 with both amplicons within one SARSeq replicate, while only 18 and 25 samples were detected twice independently with N1 or N3 amplicon, respectively. We thus conclude that SARSeq appears sensitive down to single reverse-transcribed viral genomes. We also assessed the reproducibility of SARSeq runs **(Suppl. Fig. 5A, B)**. As anticipated, the absolute read numbers are not necessarily correlated, but there is very good correspondence regarding whether or not a sample is positive. We observed the expected upper end of N1/N3 read counts produced by the end point PCR strategy to distribute sequencing space evenly across samples (**Fig. 1E**). Given the number of samples at the limit of detection, some samples were undetected in NGS but present in one qPCR replicate, and some samples were detected by NGS that were missed by one or both qPCRs. However, as mentioned above **(Fig. 5F)**, 96.3% of all samples with Ct <36 were detected in both NGS replicates.

**Figure 5.**
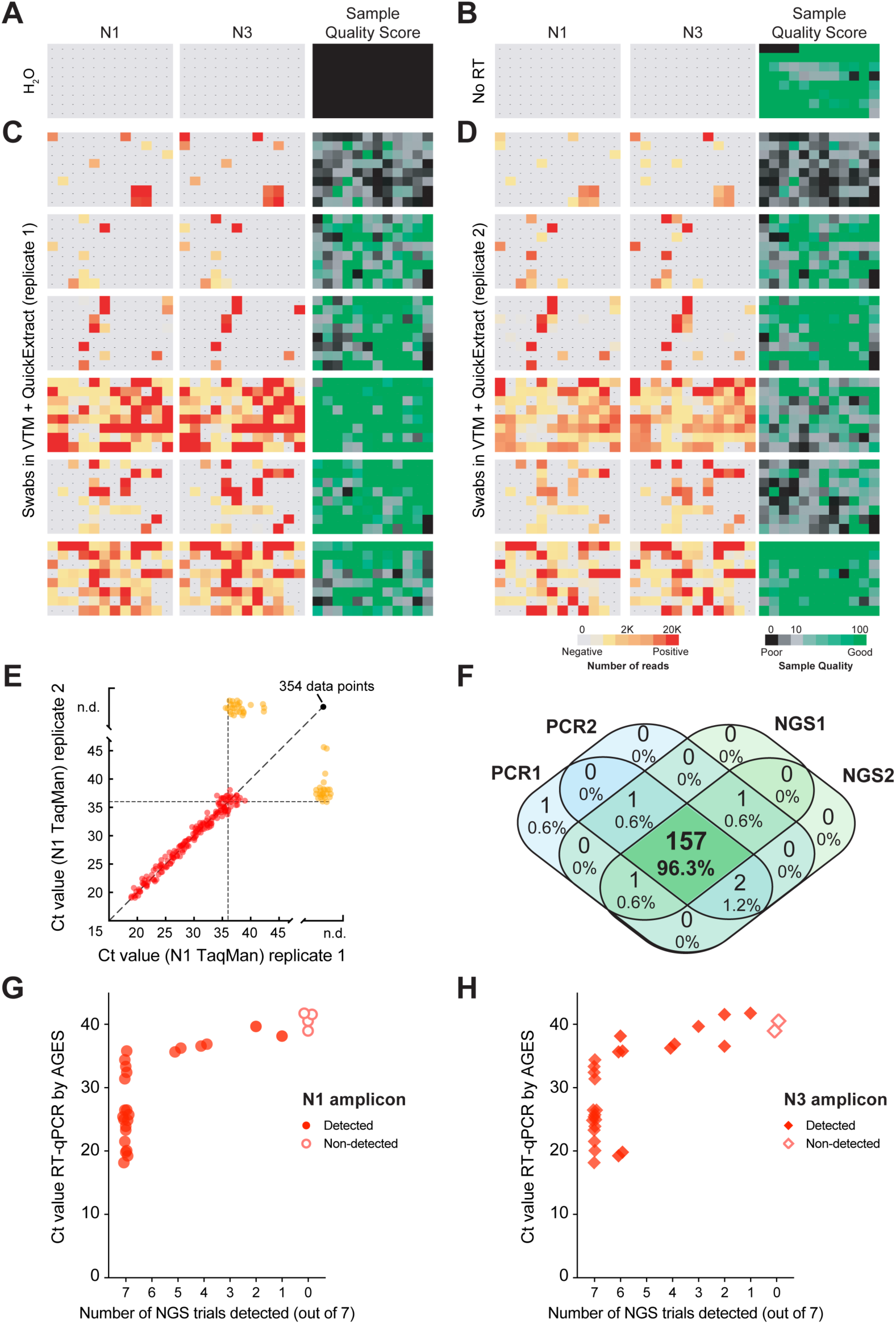
SARSeq robustly detects SARS-CoV2 in patient samples. **A**. H2O control plate of SARSeq with three primer pairs, namely N1, N3, and ribosome in the presence of RTC as well as N1 and N3 scavenger spike-ins. **B**. SARSeq on a gargle-QuickExtract plate in the absence of reverse transcriptase. No false positive wells were detected. Sample QC scores high due to the absence of RTC reads while DNA templated ribosomal amplicon is observed. **C**. Analysis of 576 samples obtained by swab and collected in VTM. **D**. Independent replicate of C based on the same inactivated patient swab. Color codes depict read counts and sample quality score for C and D. **E**. Two independent N1 TaqMan qPCR runs on samples as in C and D. Ct values of both runs are plotted against each other. Color code: red: sample scoring positive in both qPCR replicates, orange: sample scoring once, black: sample scoring negative. Stochastic and latest detection of SARS-CoV2 at cycle 36. **F**. Venn diagram of reproducibility for all samples scoring positive with Ct 36 or less for at least one qPCR replicate. **G, H**. Comparison of NGS results by SARSeq to Ct values obtained by diagnostic qPCR. A set of 90 samples (including swabs and gargle in different buffers) was used for RNA extraction and qPCR measurements and in parallel aliquots of these samples were mixed either with QuickExtract or TCEP/EDTA and measured by SARSeq in triplicates and quadruplicates, respectively. We report the number of replicates in which we called a sample positive by NGS (with the N1 or N3 amplicon) relative to the qPCR Ct values. Not shown are 63 samples that were negative by qPCR and NGS.

### SARSeq robustly detects SARS-CoV2 in samples from a human diagnostics setting

As a pilot for a systematic clinical performance study we compared SARSeq to a gold standard qPCR assay conducted in a human diagnostic setting. This test included a 90 samples of diverse nature, including swabs in VTM, gargle in isotonic NaCl, and others, of which 28 were positive according to a gold standard qPCR pipeline. This pipeline used the equivalent of 17 µl crude sample whereas we used 2.5 µl (for QuickExtract mix) and 4.7 µl (for TCEP/EDTA mix) crude original sample as input for SARSeq. We performed seven replicates for SARSeq (3 in QuickExtract and 4 in TCEP/EDTA) which yielded highly consistent results: as expected, samples that were negative by qPCR showed only 0 or 1 reads in all replicates, whereas samples that were positive by qPCR consistently displayed thousands of reads. Specifically, we detected the N1 amplicon in 7/7 replicates for all positive samples with Ct values <36.5 and in at least 1/7 replicates for all others with Ct values <38.9 (**Fig. 5G)**. The N3 amplicon showed a similar pattern but seems more sensitive to sample quality (**Fig. 5 H**). We also assessed quantitativeness by comparing Ct values from the qPCR directly to the read counts obtained by NGS (**Suppl. Fig. 5C, D**). As intended by the endpoint PCR for dynamic-range compression, SARSeq is blunted for high viral titers (Ct values lower than ∼33) but is semi-quantitative for weakly positive samples. We therefore conclude that SARSeq is a useful method to detect SARS-CoV2 in clinical and diagnostic settings for samples of various chemical compositions, robustly detecting samples with Ct values ∼36, but also samples up to Ct values of 39, albeit with decreasing probabilities.

### SARSeq can detect multiple respiratory viruses in a single reaction

Multiple infectious agents cause diseases with overlapping clinical symptoms to COVID-19, including influenza A and B virus, parainfluenza virus, rhinoviruses, and respiratory syncytial virus. It is expected that, particularly in the winter season, various respiratory symptoms will cause concerns and thereby dramatically increase the demand for SARS-CoV2 tests. For SARSeq, adding amplicons corresponding to other infectious agents comes at little extra cost as long as it does not increase the required sequencing depth. Therefore, we can further multiplex SARSeq to detect other common respiratory viruses (or other pathogens) found in the same sample used for SARS-CoV2 testing.

As proof of principle, we optimized primers for influenza A virus, influenza B virus, and rhinovirus to be combined with our SARS-CoV2 specific SARSeq pipeline. To this end we selected primers based on qPCR performance, amplicon length, and an NGS pilot experiment. For a pan-influenza A amplicon we settled on combining a degenerated forward primer from Bose et al. with a degenerate WHO reverse primer, both targeting the M gene^37,38^. For pan-influenza B, we selected a previosuly characterized primer pair binding to the M gene^38^. Rhinovirus was detected using a primer pair described previously^39^.

To test performance across a large number of specimens, we used sample plates from gargle collected in HBSS via our in-house testing pipeline (negative for SARS-CoV2) and spiked in purified RNA obtained from HEK293T cells infected with respective virus strains at a ratio of 1:100 (per gargle volume) or dilutions thereof. Samples were processed using the protocol and robotic pipeline as for other experiments, except that we performed PCR 1 in the presence of six primer pairs, two against SARS-CoV2 (N1 and N3), and one each for ribosomal control, influenza A, influenza B, and rhinovirus. Upon pooling of 12 samples each and PCR 2, samples were sequenced and reads were mapped back to individual wells (**Fig. 6A**). Of note, by pipetting the viral RNAs prior to setting up the RT reaction we apparently generated a contamination of influenza B RNA that was stochastically distributed across all reactions (including H_2_O controls but not -RT controls) and obtained reproducibly lower read counts than the samples to which we intentionally added this viral RNA (**Fig. 6B**). This event emphasizes the importance of complete local separation of reaction mix preparation and sample handling. All post-PCR steps were always performed in a different lab from the initial setup of the reaction and thus DNA amplicons did not generate detectable contamination of our pipeline. Nevertheless, we implemented incorporation of UTP and a UDG digestion step prior to PCR in our protocol to protect against contamination risk with DNA amplicons (see Methods).

**Figure 6.**
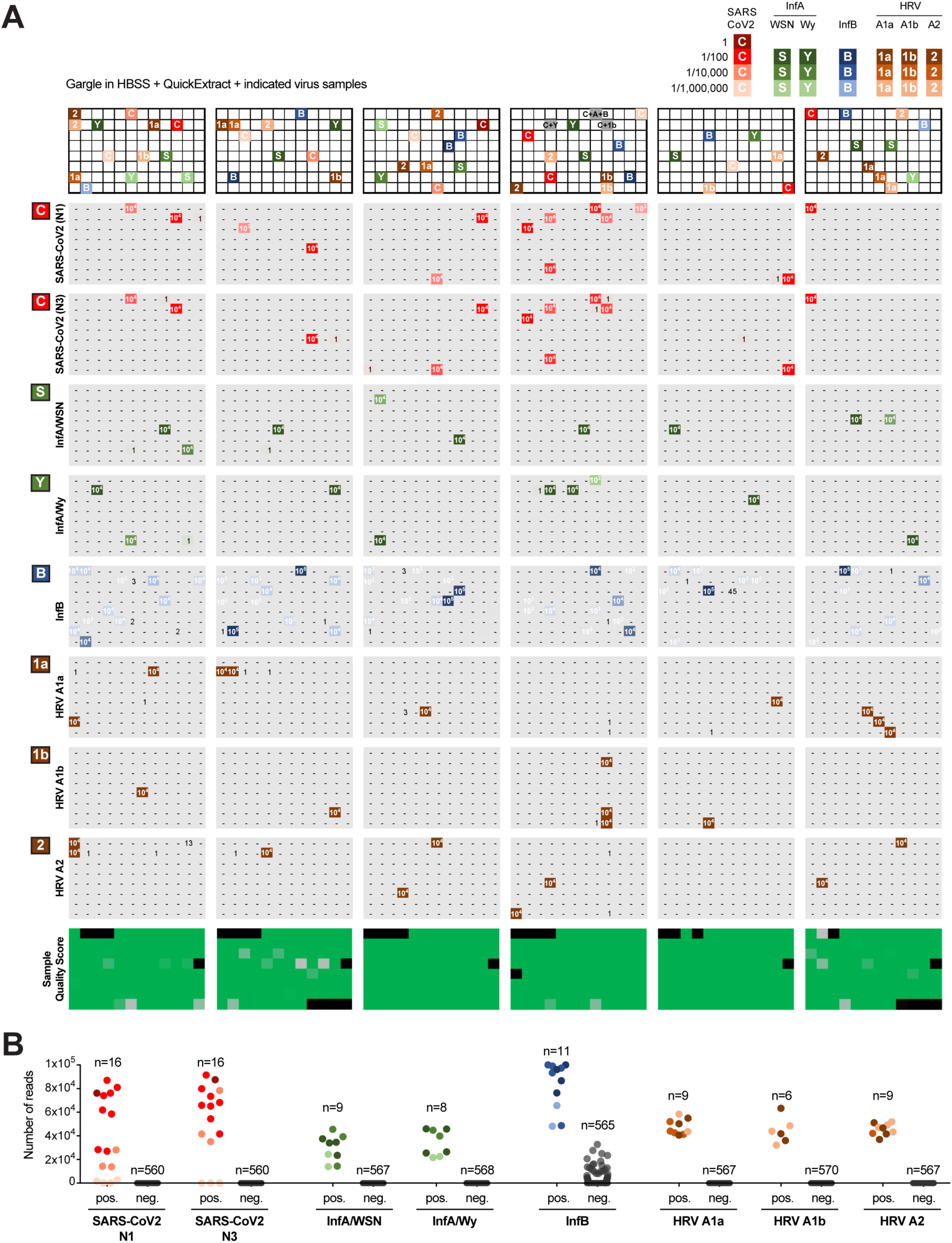
SARSeq can detect multiple respiratory viruses in a single reaction. **A**. Six 96 well plates filled with gargle in HBSS and inactivated in QuickExtract from our in-house pipeline were spiked with RNA from various respiratory viruses. For SARS-CoV2, a positive gargle sample with Ct value of 30 was diluted as indicated. RNA for all other viruses was obtained from HEK cells 48 h after infection with the virus. Dilution indicated by voluminometric ratio. SARSeq was performed with six primer pairs in one reaction, namely N1 and N3 for SARS-CoV2, Influenza A virus, Influenza B virus, human rhinovirus (HRV), and the ribosome. Influenza A substrains and HRV substrains were distinguished based on amplicon sequence variants. **B**. Analysis of false positive and false negative rate of the experiment in panel A. As expected, 1:100,000 dilutions of the SARS-CoV2 sample with a Ct of 30 were missed. All other positive samples were detected for all viral spike-ins. False positive samples were detected for Influenza B virus presumably due to sporadic contamination of reagents or equipment with purified RNA also in the H20 control plate (not shown). False positive read counts did not reach numbers observed in truly positive samples. No false positive samples above threshold were detected for any other virus.

Our multiplexed pipeline to detect RNA of various viral respiratory diseases in parallel performed robustly across six 96-well plates. We obtained no false negative samples besides, as expected, the 1/1mio dilutions of a SARS-CoV2 sample with Ct=30. Aside from the apparent influenza B contamination, we also did not see any other false positives. Moreover, the amplicon sequence of influenza A virus allowed us to distinguish between two different sub-strains we had used, namely A/Wy and A/WSN **(Fig. 6B)** the latter of which is anticipated to circulate in the northern hemisphere 2020/21 flu season^40^. Similarly, with a single primer pair we were able to distinguish between the three rhinoviral strains, namely HRV A1a, A1b, and A2 by polymorphisms in respective amplicons. Taken together, our pipeline in its current form differentially detects seven different viral respiratory agents in a single reaction, contains various internal controls, a sample quality control, and by design has particular sensitivity for SARS-CoV2. SARSeq thus represents a multiplexed, massively parallelized assay for *saliva analysis by RNA sequencing* to detect respiratory infections by means of RT-PCR and NGS.

## Discussion

It has been proposed that mass testing for SARS-CoV2, with a focus on surveillance of asymptomatic individuals, can help mitigate the effects of the COVID-19 pandemic, and this strategy has shown good results in South Korea, Taiwan and Singapore^7,9^. To further expand this to other nations, the assays used for such large-scale testing must meet a number technical considerations: i) the tests themselves must be highly specific to avoid false positives leading to the isolation of individuals based on erroneous results; ii) the costs for mass testing must be as low as possible to reasonably enable scaling; iii) the assay must be massively scalable and return results in a short timeframe; iv) mass testing must not interfere with testing in medical/diagnostic facilities, it is thus preferable that it is neither carried out at the same facilities nor competes for supplies required to diagnose symptomatic patients.

Here, we described SARSeq, an NGS-based testing method that meets the technical considerations outlined above. The current design enables analysis of up to 36,000 samples in parallel, and we demonstrated the analysis of >18,000 samples in a single sequencing run. SARSeq shows high specificity at two levels: at the amplicon level it has maximal specificity as it detects the precise sequence of two independent SARS-CoV2 amplicons; at the sample identification level, SARSeq employs a number of measures to completely suppress mis-assignment between samples. Moreover, the cost per sample analyzed by SARSeq is low compared to other available tests; it relies on common reagents and enzymes that can be purchased at scale or produced in-house with standard biochemical methods. The costs of sequencing per sample also become negligible considering that they are divided over thousands of samples. Finally, SARSeq, with the exception of NGS, relies exclusively on equipment available in most molecular biology facilities in academia and industry, and does not compete for resources with other diagnostic tests.

Several alternative methods to detect SARS-CoV2 by NGS have been developed. While some focus on viral genome sequencing and are thus of lower throughput^20,41^, others aim to be used for detection of viral infection by amplicon sequencing at high throughput similar to SARSeq. In one strategy, samples are indexed during the RT step and PCR is performed in pool^19^. This approach has the advantage that early sample pooling circumvents the need for large numbers of individual PCR reactions. We anticipate however, that such an approach would maintain the vast dynamic range in viral titers between samples. As explained above, this leads to highly positive samples dominating the available NGS read space, thereby prohibiting true scalability while maintaining sensitivity.

In contrast, SwabSeq and SARSeq use individual PCR reactions for each sample, which allows dynamic-range compression by end-point PCR. In addition, both methods use a dual indexing strategy to gain the required robustness in sample recall that is key for diagnostic assays. However, SwabSeq and SARSeq differ in several important aspects that directly impact scalability and the multiplexed detection of different amplicons:

SwabSeq uses a 1-step RT-PCR reaction, whereas SARSeq performs RT and PCR in two steps, which we found to significantly suppress unspecific amplicons and thus make efficient use of the read space, a prerequisite for sensitivity, scalability and multiplexing of amplicons. However, a one-step RT-PCR reaction is also compatible with SARSeq, albeit with slightly lower amplicon specificity (Fig. 1D).

More importantly, due to a single indexing step, upscaling of Swab Seq is l inear rather than combinatorial – every additional sample requires one additional primer pair per amplicon. In contrast, SARSeq uses a two-dimensional indexing strategy, which allows combinatorial (multiplicative) scaling of up to tens of thousands of samples with just a few hundred primer pairs and therefore leverages the sequencing capacities offered by NGS. By linear indexing, such dimensions would neither be cost effective nor logistically feasible.

Another important difference is that the amplicons generated by SwabSeq contain the flow cell adaptors but not the i5/i7 sequencing primer bindings sites and thus require a mix of custom sequencing primers, one for each amplicon. This limits the number of different amplicons that can be surveyed in parallel and also can cause heterogeneity in cluster signal intensity and thus impact sequencing quality. In contrast, all SARSeq amplicons contain standard i5/i7 sequencing primer bindings sites and are directly compatible with the regular sequencing protocols and reagents on all Illumina platforms. This facilitates the addition of further amplicons to the assay and since all are read out by the same standard i5/i7 binding sequencing primers. Therefore the extra PCR reaction that SARSeq requires, is small technical burden, which however allows dramatic improvements in scalibity of samples and amplicons and yields amplicons with homogenous sequencing properties.

Together, these advantages allowed the detection of other respiratory RNA viruses beyond SARS-CoV2. Specifically, we used SARSeq to detect influenza A virus, influenza B virus, and human rhinovirus and this list can be easily expanded to additional infectious agents, both circulating and also newly emerging pathogens. Moreover, SARSeq is not limited to respiratory specimens, but we envision that this pipeline could be used for other human and animal samples or even monitoring pipelines such as those that sample wastewater.

SARSeq solves the technical challenges downstream of sample acquisition, shifting the bottleneck towards truly high-scale testing from the actual assay to developing matching logistics for sample collection, maintaining supply chains, developing appropriate data management tools, and also often overcoming legal hurdles. However, we envision ways in which SARSeq can be implemented right away, to already significantly contribute to detecting infection events before they spread. In the first, samples can be collected and inactivated locally, potentially also first PCR might be performed using prepared and distributed primer arrays, then shipment to centralized location for PCR2, sequencing and analysis. Importantly, depending on the legal situation and aim, this pipeline does not necessarily need to a human diagnostics lab and would thus not block important infrastructure. In the second, companies, universities, other types of institutions could implement a regular sample collection strategy among employees/students/other members and team up with a local academic or industry lab that can with relatively little effort implement this protocol.

Different SARS-CoV2 detection assays have been optimized over the last few months, each with strengths and limitations. For SARSeq, a potential limitation is the time requirement of the assay. Two PCR reactions must be performed followed by NGS and analysis, so the theoretical minimum time required is around 15 hours. In practice, our tests took at least 24 hours from sample preparation to results. Therefore, SARSeq is not ideally suited for situations where immediate results are required. In such cases, antigen tests^42^ or RT-LAMP^43,44^ are superior methods. Rather, SARSeq is ideally suited for regular (e.g., once or twice a week) surveillance of infections in a large scale population, with high sensitivity and specificity (i.e. negligible false positive and false negative rates). SARSeq might also be suitable to test symptomatic persons if a turnaround of 24h for the test itself is acceptable. In addition, SARSeq can be implemented in epidemiology studies to understand the spreading dynamics of infections^45^ and to investigate interaction between different pathogens across large populations^46^. However, the main advantage of SARSeq is that the same turnaround time of 15-24 hours can be used to simultaneously test tens of thousands of samples. Therefore, SARSeq complements available diagnostic tests, increasing capacity to enable large-scale monitoring efforts.

## Supporting information

Suppl. Table 1 - Primer sequences

Suppl. Table 2 - Pipetting scheme

## Data Availability

The analysis script is available on GitHub at https://github.com/alex-stark-imp/SARSeq and at
https://starklab.org.

## Author Contributions

R.Y., L.C. and U. E. performed experiments with help of E.Ö. and M.S.. Experimental design was by R.Y., A.S., L.C. and U. E.. All NGS analysis was performed by A.S.. A.B. set up and performed automated pipetting. A.V. supported optimization of RT and PCR conditions, oversaw and executed NGS. R.H. shared reagents, expertise, and supported experimental design. K.U, B.H., and D.K. purified proteins used in this study. J.B. cloned various constructs and provided technical help. E.S., A.K-K. and S.I provided clinical samples, J.S., P.H. and F.A. provided samples and results from a human diagnostics lab. The VCDI is a consortium of multiple people that established various SARS-CoV2 testing pipelines in close collaboration and supported sample acquisition as well as multiple steps of optimizations and upscaling. F.A. oversaw the human diagnostics pipeline, assessed results and corrected the manuscript. Figures were generated by L.C. and U.E.. A.S., L.C., and U.E. supervised the study, and wrote the manuscript.

## Acknowledgements

In immediate response to the start of the pandemic, many people in our campus united to contribute with their skills in molecular biology towards the developing of various testing strategies. This task force developed bottom up with contributions from people at all levels and diverse expertise, and it came to fruition with a robust qPCR-based pipeline for in-house testing of employees and other entities (led by Johannes Zuber and Stefan Ameres among others), the development of an improved RT-LAMP test (led by Andrea Pauli and Julius Brennecke), as well as the NGS method presented in this manuscript. In addition, strong alliances with public health authorities and politicians had to be formed. These acknowledgements inevitably fall short in thanking everyone involved, but we wish to highlight those who made the biggest contributions. Harald Isemann coordinated the strong administrative and financial support from our institutes to these endeavors, for which we are extremely grateful. We thank the entire RT-LAMP team for their close interaction and support in various aspects, in particular Andrea Pauli and Julius Brennecke for their tremendous help in obtaining patient samples. Various people invested their time and effort to implement the in-house testing pipeline that enabled us to safely come back to work (Johannes Zuber, Harald Scheuch, Peter Steinlein, Stefan Ameres, Sabina Kula), and also provided samples for this work (Katharina Bergauer, Martina Weissenböck, and Barbara Werner). We are extremely grateful to Irma Salimović-Bešić, and Sandra Vegar-Zubovic for providing patient samples in difficult days. We are indebted to Tim Clausen and Anton Meinhart for supporting enzyme purification efforts, to Andreas Sommer, Ido Tamir and the entire NGS team at the VBCF. We also thank Raphael Manzenreither for technical help, Gijs Versteeg for virus preparations and other help, and Edith Soucek for support. We thank Katrina Woolcock from Life Science Editors for editing, as well as the community of COVID fighters worldwide. We thank all employees for consent to sample use and ethics boards for rapid processing of the requests. Last but not least we would also like to thank our labs and all coworkers for understanding and supporting us in so many ways. R.Y. was partially supported by Oliver Bell’s laboratory at USC. Curiosity driven biomedical research at the IMP is largely sponsored by Boehringer Ingelheim. IMBA is generously funded by the OEAW.

The current pandemic has challenged healthcare systems worldwide, but it has also lead to an unprecedented concerted response on multiple levels. We wish to contribute to the fight and therefore welcome anyone interested in implementing this method to contact us. Further developments and answers to reagent requests will be handled through www.sarseq.org (in preparation) to enable high-throughput testing across all our communities.

## Supplemental Figures

**Suppl. Fig. 1.**
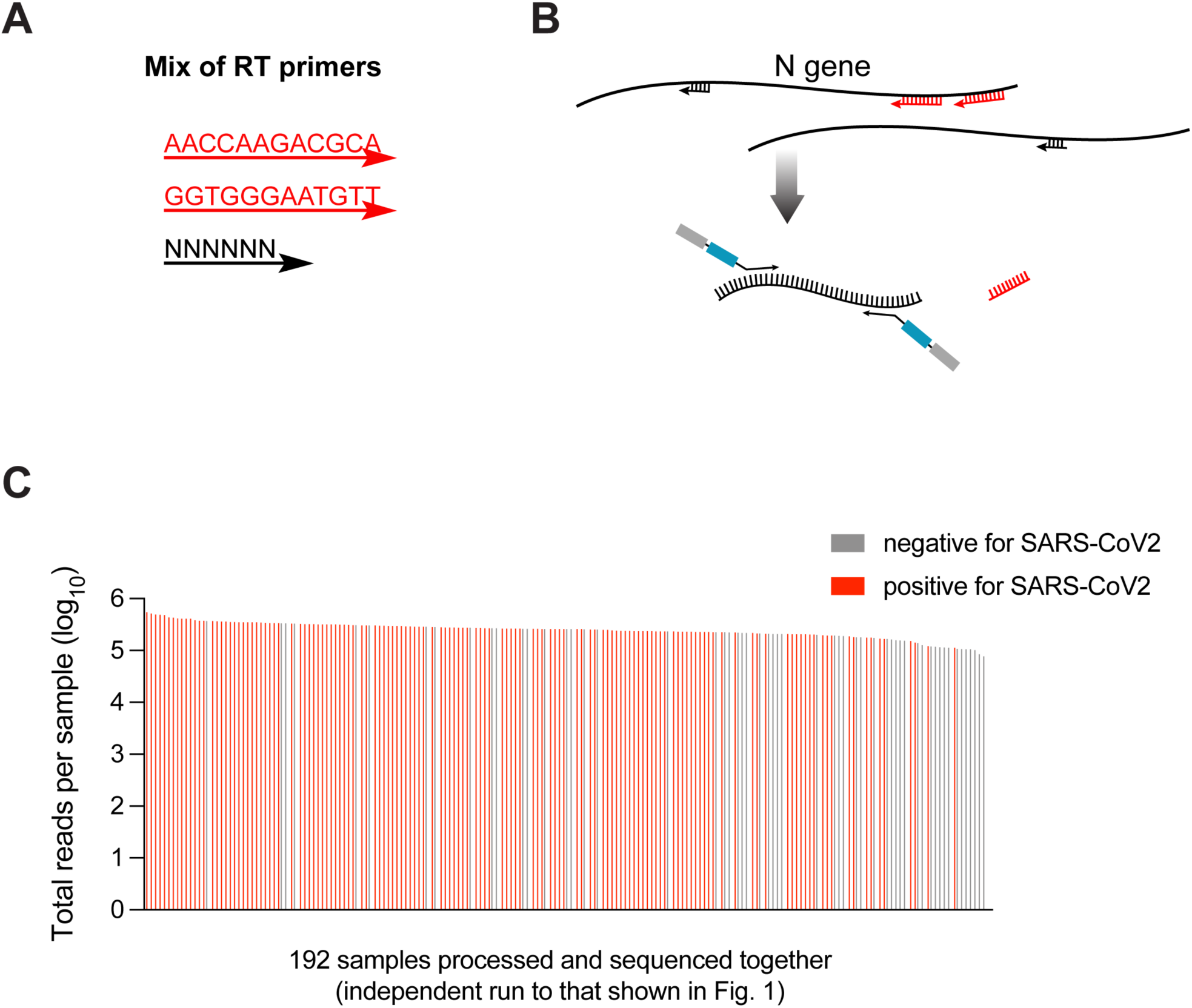
Two-step RT-PCR strategy enables specificity and even read distribution. **A**. The RT primer mix contains two N-gene specific 12-mers (actual sequences shown) in addition to random hexamers. **B**. Scheme of the RT priming sites relative to the specific PCR primers. For SARS-CoV2 the 12-mers generate a cDNA into which the PCR primers are nested. **C**. Total reads per sample across a set of 192 samples that contain several negative samples as well as positive samples with titers spanning 4 orders of magnitude. Our 2-step RT-PCR strategy followed by end-point PCR lead to a very even distribution of NGS reads per sample independent of viral titer, ensuring equal representation on the sequencing flow cell.

**Suppl. Fig. 2.**
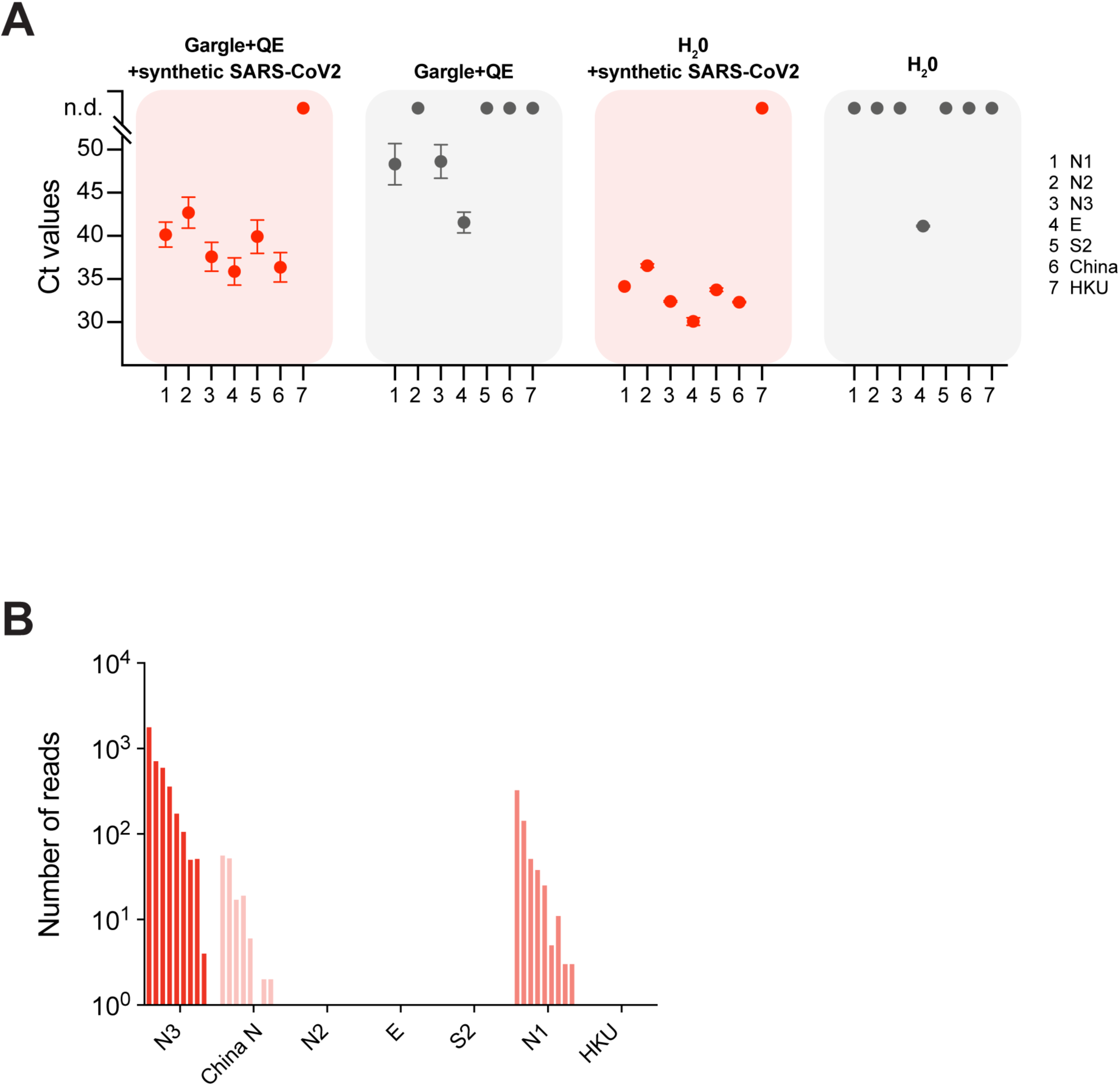
Choice of SARS-CoV2 specific primers. **A**. SYBR green based qPCR on samples generated by mixing gargle from a healthy individual or water, with 1000 copies of synthetic SARS-CoV2 RNA. Primers used were based on previous publications^28,47^, but were extended at the 5’ by the sequences required for NGS, namely i5/i7, a random stagger, and a barcode. Several primer pairs showed a good template dependent generation of double stranded DNA. **B**. A next generation sequencing pilot showed good amplification for N1 and N3, thus we chose N1 and N3 amplicons to detect SARS-CoV2 in the presented setup.

**Suppl. Fig. 3.**
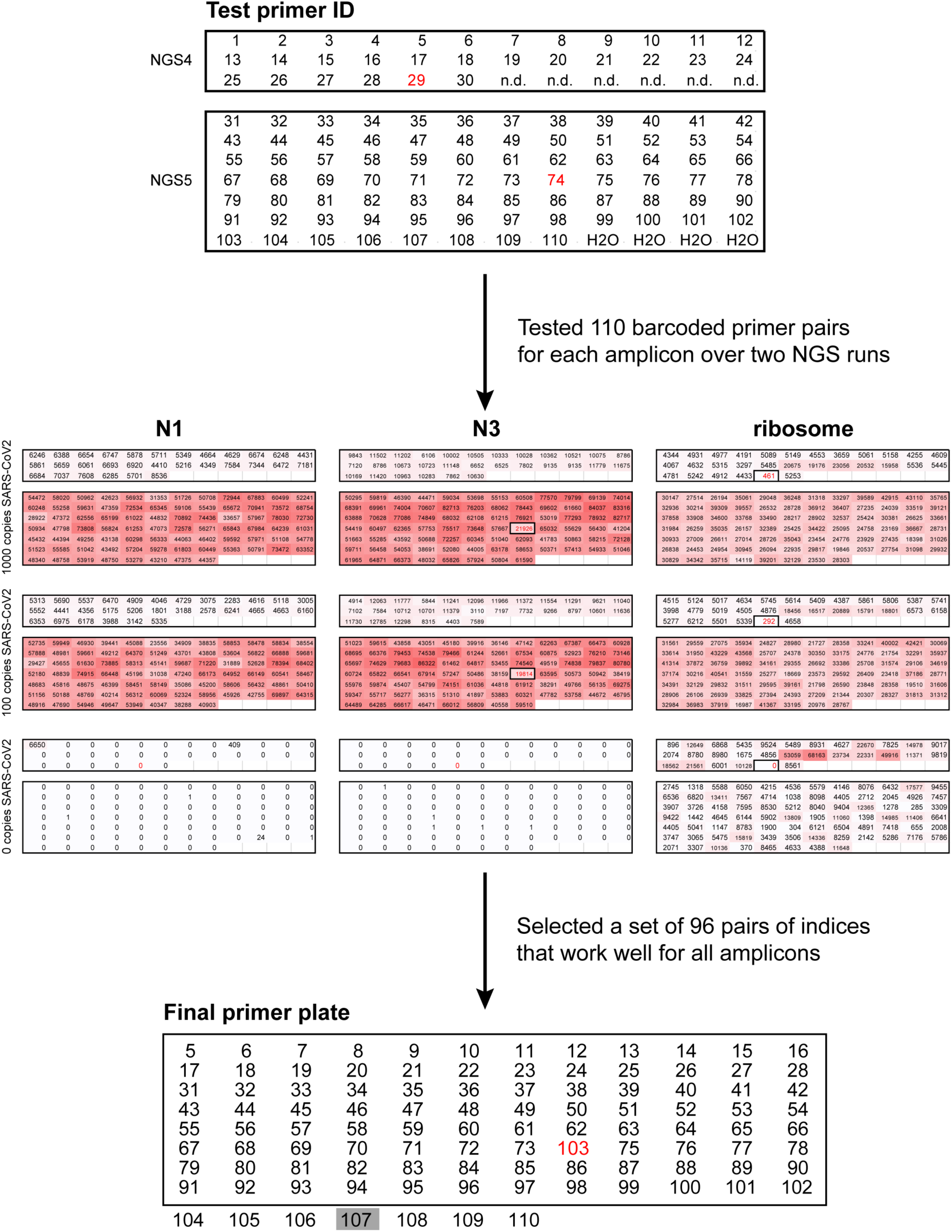
Validation of the amplicon-specific primer pairs containing 96 unique dual indices. Testing of primer pairs for three amplicons (SARS-CoV2 N1 and N3, and human ribosome) carrying extensions with 110 unique dual indices. Primer pairs with indices #29 were excluded as they did not efficiently produce the ribosome amplicon; primer pairs with indices #74 were excluded as they did not efficiently produce the N3 amplicon. Primer pairs 1-4 can be used efficiently but were not included in the final set because these indices were frequently used in the lab during initial setup of the method and we wanted to eliminate all possible sources of cross contamination.

**Suppl. Fig. 4.**
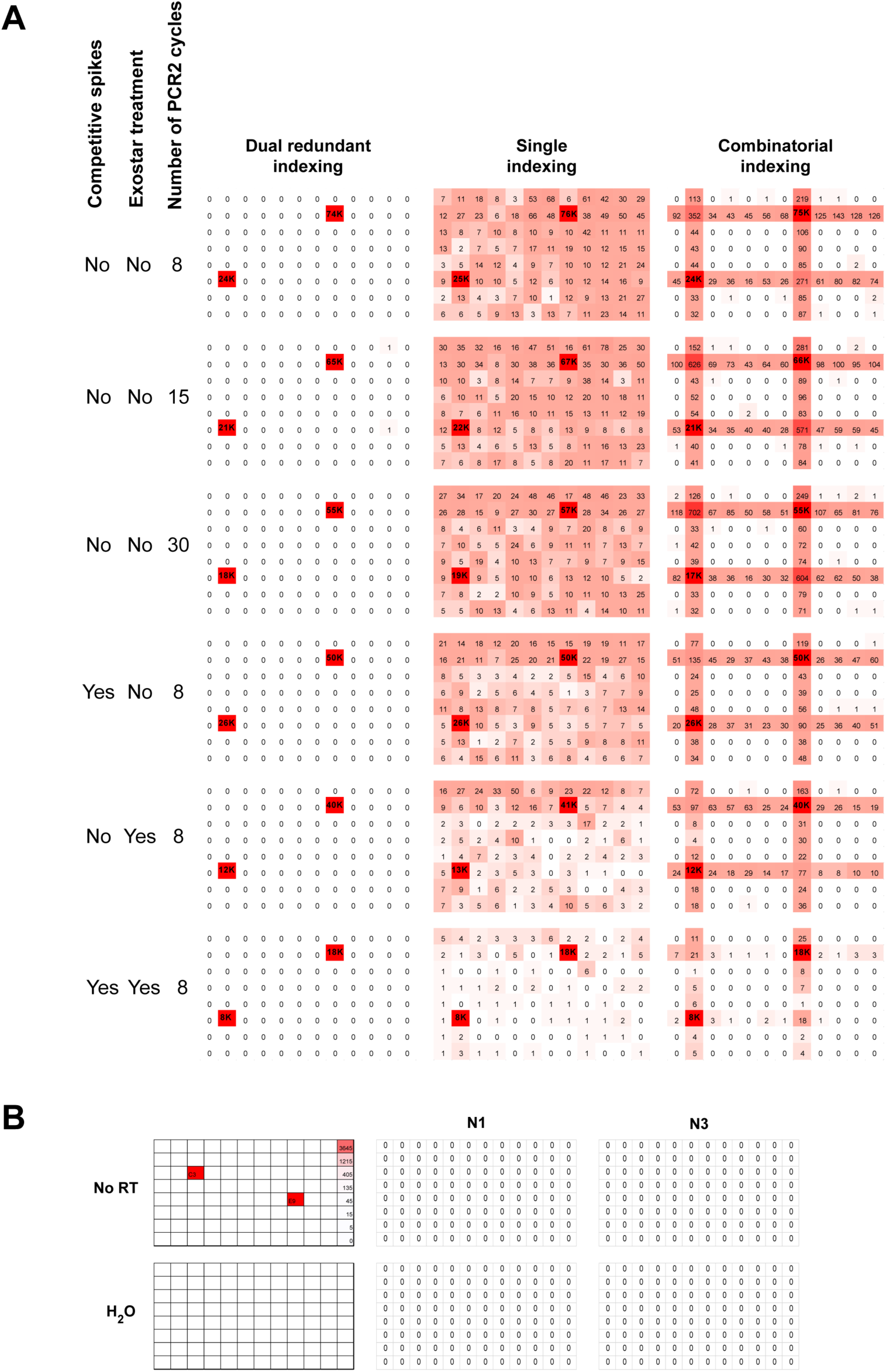
**A**. Contribution of the different measures taken to completely suppress mis-assignment of indices to incorrect samples (and in that way avoid false positives). SARS-CoV2 synthetic RNA was added to positions B8 and F2. Assigning sample identities based on single indices or combinatorial indices produces a substantial amount of misassignments. These can be reduced with three independent measures: addition of a competitive spike-in and treatment with Exostar to remove any left-over primers after PCR1, and limiting the number of cycles in PCR2 to prevent recombination between amplicons and their indices. Whereas all these measures help reduce misassignments, adding dual redundant indices (or unique dual indices) completely suppresses this. **B**. Shown are N1 and N3 reads for two negative control plates (-RT and H_2_O) that were run together with numerous other positive plates in the same run. The fact that we detect 0 reads shows that there is no misassignment of well identities across plates either. This is because each plate is also encoded by redundant/unique dual indices.

**Suppl. Fig. 5.**
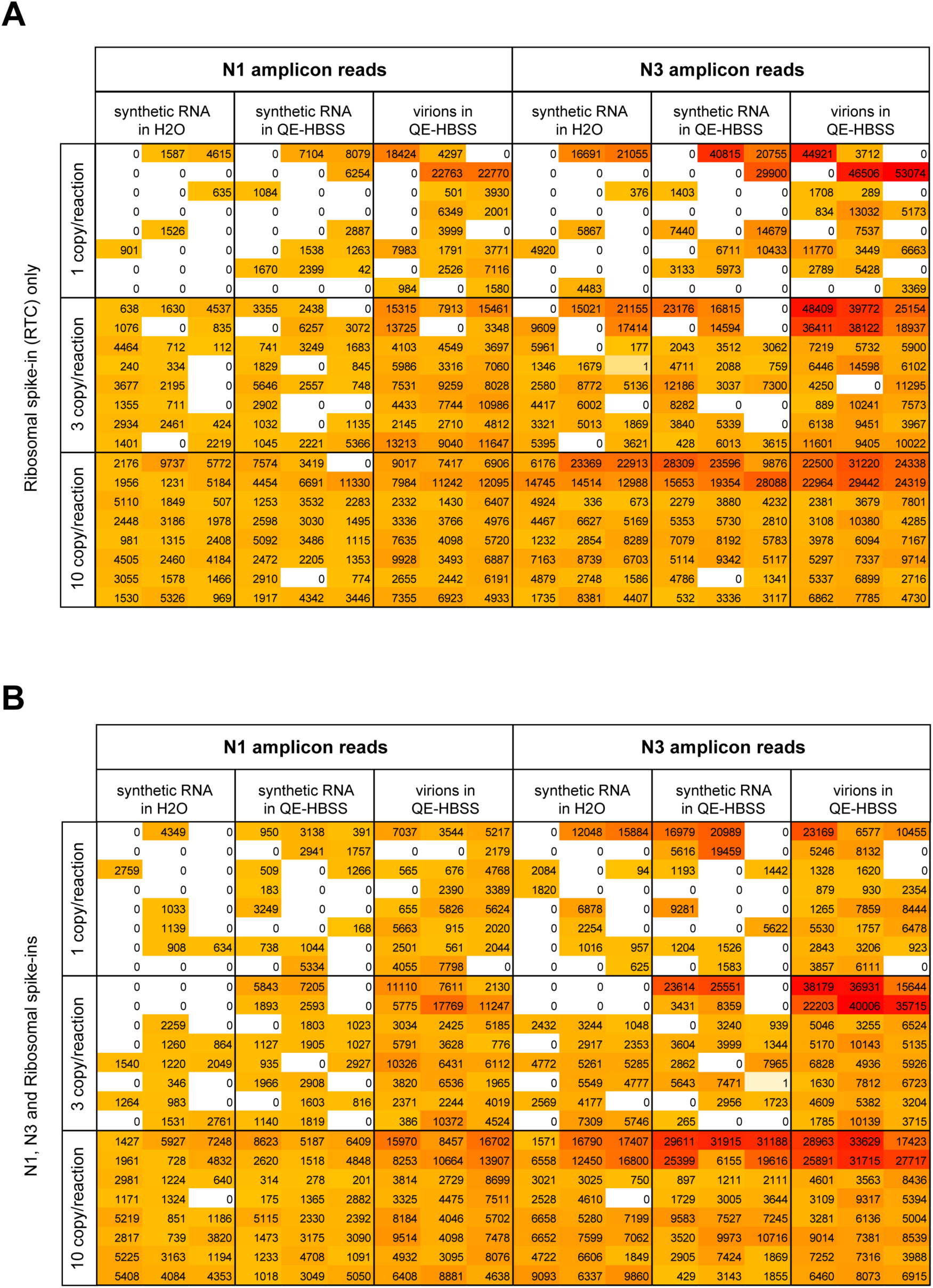
Raw data for limit of detection experiments presented in Fig. 4B. **A**. Read count table obtained in absence of N1- and N3-control-spike-in. **B**. Read count table obtained in presence of N1- and N3-control-spike-in.

**Suppl. Fig. 6.**
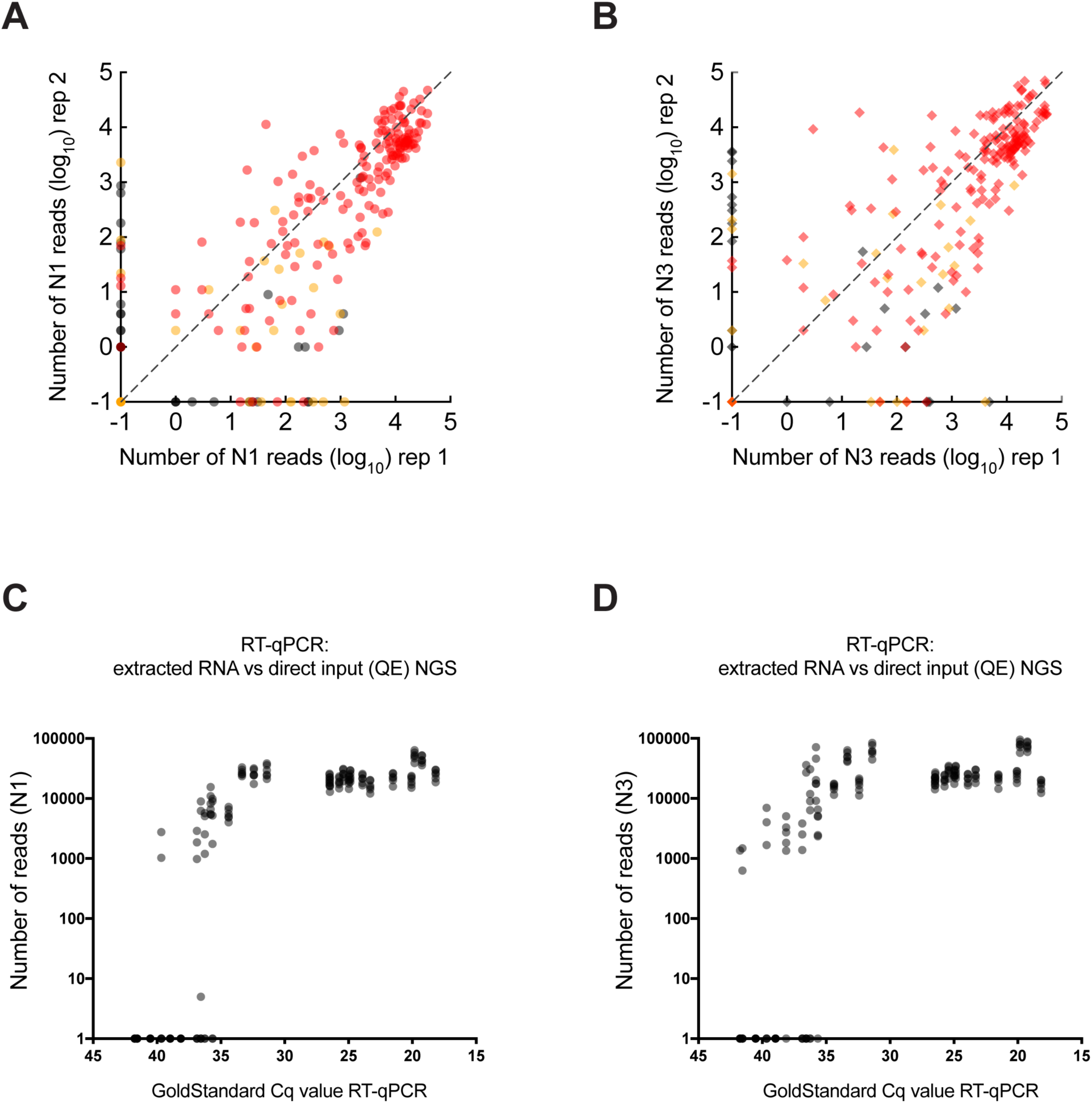
Semiquantitative behaviour of SARSeq on patient samples. **A, B**. Correlation of read counts of N1 (A) and N3 (B) amplicons across two independent SARSeq runs of the samples shown in Figure 5C, D. In red are individual samples also detected in two qPCR replicates, in orange are samples detected in one out of two qPCR replicates, and in gray those that were not detected by qPCR. **C, D**. Correlation between read counts for N1 (C) and N3 (D) amplicons and Ct values obtained by diagnostic qPCR. The qPCR analysis was performed on purified RNA, in parallel crude samples were measured in seven replicates by SARSeq, all replicates are shown as individual circles. SARSeq is robust until ∼Ct 36 and becomes probabilistic in samples with lower viral titers.

## Materials and Methods

### Sample material and ethics

The present study includes preliminary investigations and results of a clinical performance study approved by the local Ethic Committee of Vienna (#EK 20-208-0920). For that, left-over samples from healthy participants were obtained from an anonymous routine SARS-CoV2 screening pipeline, and left-over patient samples in Fig. 5G,H were obtained by the Austrian Agency for Health and Food Safety (AGES) in a diagnostic pipeline and provided to us fully anonymized. For VTM samples used for Figure 5 A-F, an additional approval (#06-04-9-33163 from 21/07/2020) was obtained from the Ethics Committee of the Clinical Center of the University of Sarajevo. The study was conducted in accordance with the Declaration of Helsinki.

### Input sample preparation

The pipeline we describe can start from a variety of different input samples. The types of samples we have tried are:

- Purified RNA from gargle samples
- Gargle samples mixed with DNA QuickExtract solution from Lucigen (1:1 ratio)
- Swabs in VTM mixed with DNA QuickExtract (1:1 ratio)

Samples mixed directly with QuickExtract were incubated for 5 min at 95°C for inactivation and used directly or stored frozen at −80°C. We did not observe a decline in positive signals upon freezing and thawing (even after two cycles of freeze-thaw).

The samples were arrayed in 96-well plates. The described reaction setup uses up to 5 µL of any of the above described samples.

### Reverse transcription

Reverse transcription was performed with reverse transcriptase, homemade Ribonuclease inhibitor and a primer mix containing random hexamers as well as two 12-mer oligonucleotides that prime on the SARS-CoV2 N gene.

A master mix containing all components listed below was prepared and distributed to 96-well plates (20 µL per well). Using a liquid-handling robot (or multi-channel pipettes), 5 µL of each sample were transferred to each individual well containing the RT reaction mix. RT reactions were set up at room temperature. Plates were sealed with aluminum sealing foil (facilitates easy removal after RT reaction that reduces vibrations in wells avoiding generation of aerosols which may cause cross contamination between samples) and incubated in a thermocycler following conditions listed below.

#### Master mix composition per reaction/well (volumes in µL)

|  |  |
| --- | --- |
| 10× RT Buffer<sup>A</sup> | 2.5 |
| 25 mM each dNTP | 0.5 |
| 1M DTT | 0.1 |
| RT primer mix<sup>B</sup> | 2.0 |
| Ribonuclease inhibitor | 0.5 |
| Reverse transcriptase | 0.5 |
| Water | 13.8 |

Wherever mentioned, for each reaction 1000 copies of Ribosome synthetic RNA spike-in and 50 copies of each N1 and N3 RNA synthetic spike-in were included in the RT reaction master mix. Also, wherever mentioned Thermofisher/Invitrogen™ SuperScript™ III or Luna Universal One-Step RT-qPCR Kit (NEB) or homemade reverse transcriptase 2.5 (see below for details) was used for reverse transcription. In all other experiments, homemade reverse transcriptase 3 was used for reverse transcription.

#### Thermocycler program

5 min at 25°C (primer annealing)

15 min at 55°C (reverse transcription/RT was carried out at 42°C for reverse transcriptase 2.5) 3 min at 95°C (RT inactivation)

Cool down to 12°C (removing the plate while it is still hot will cause bending of the plastic, making further pipetting and sealing more difficult)

#### ^A^10× RT Buffer composition

200 mM Tris-HCl pH 8.3

500 mM KCl

50 mM MgCl_2_

200 mM (NH_4_)_2_SO_4_

1% Triton X-100

#### ^B^RT primer mix composition

12.5 mM of each random hexamer, N gene specific 12-mer #1 and N gene specific 12-mer #2 (final concentration in the complete 25 µL RT reaction is 1 mM each)

#### *In-vitro* transcription of spike-in synthetic RNA templates for reverse transcription controls

For Reverse Transcription Control (RTC), gBlock was obtained from IDT. Using IDT synthetic template RTC was PCR amplified and cloned into pCR2.1 plasmid by TOPO cloning. For cloning N1 spike-in N1FF, N1FR, N1FR, N1RR, insertF and insertR, oligos were annealed and cloned into pCR2.1 plasmid by ligation at SpeI and EcoRI sites. Similarly, for cloning N3 spike-in N3FF, N3RF, N3FR, N3RR, insertF and insertR, oligos were annealed and cloned into pCR2.1 plasmid by ligation at SpeI and EcoRI sites. RTC gBlock sequence and oligo sequences used to clone N1 and N3 spike-in templates are given below in table. Spike-in template containing plasmid clones are confirmed with Sanger sequencing. For efficient in-vitro transcription, plasmids were linearized downstream of the T7 promoter and spike-in template by cutting with a unique restriction enzyme. *In-vitro* transcription was carried out using NEB HiScribe™ kit according to manufacturer’s instructions. Transcribed reactions were treated with Turbo DNAse/ Thermofisher for 1 hr and RNA is purified using Zymo RNA clean and concentrator spin columns. RNA was aliquoted and stored at −80°C.

**Master mix composition per reaction/well (volumes in µL)**

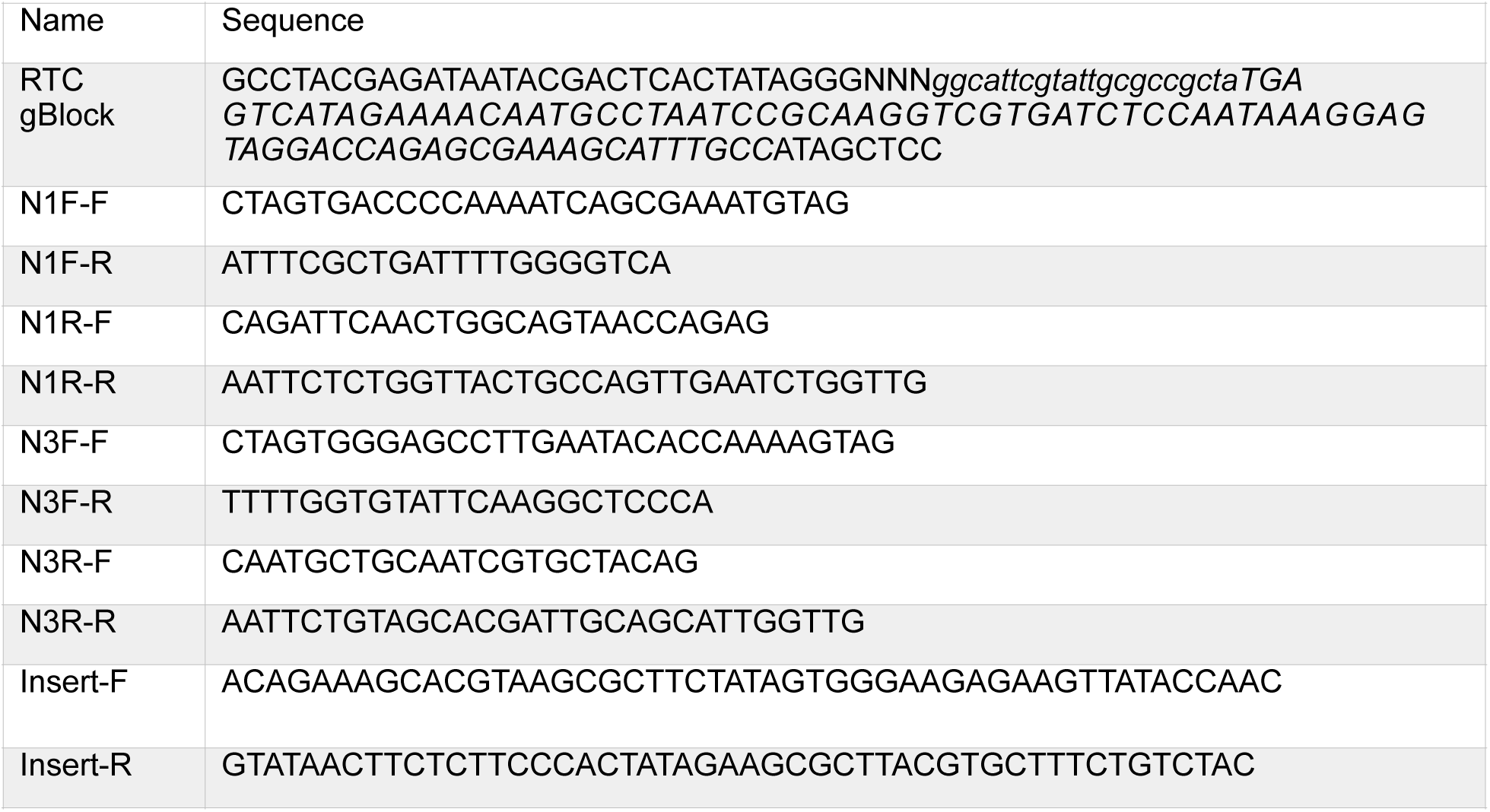

### First PCR (sample indexing)

A master mix containing all components listed below, including homemade HotStart Taq Polymerase and Uracil DNA glycosylase (Antarctic Thermolabile UDG from NEB) was prepared and distributed to a deep-well 96-well plate. The 96-primer pair combinations^C^ containing dual well barcodes were also arrayed in 96-well plates (multiple primer plates can be prepared simultaneously and stored frozen at −20°C). Using a liquid-handling robot, the 96 sets of barcoded primers were added to the PCR master mix and mixed thoroughly. 25 µL of this complete 2× PCR mix were added to the 25 µL RT reactions prepared as above.

Plates were sealed with aluminum sealing foil and incubated in a thermocycler following the conditions listed below.

All components were kept at room temperature during reaction set up; together with the first step in the thermocycler, a 10 min incubation at 30°C, this provides the right conditions for UDG to act on Uracil-containing amplification products of previous PCR reactions, thereby removing spurious carry over contaminants. After UDG heat inactivation, the subsequent PCR reaction was again carried out in the presence of UTP to prevent carry over contamination in following runs.

#### Master mix composition per reaction/well (volumes in µL)

**Master mix composition per reaction/well (volumes in $\mu\text{L}$ )**
|  |  |
| --- | --- |
| 10× PCR top up Buffer<sup>D</sup> | 2.5 |
| 100 mM dUTP | 0.07 |
| Hotstart Taq Polymerase | 0.5 |
| Antarctic thermolabile UDG | 0.5 |
| Water | 16.43 |

#### Thermocycler program

10 min at 30°C (for high UDG activity)

3 min at 95°C (UDG inactivation and Hotstart Taq activation)

45 cycles of: 20 sec at 95°C, 30 sec at 58°C, 20 sec at 72°C

2 min at 72°C

Cool down to 12°C (removing the plate while it is still hot will cause bending of the plastic, making further pipetting and sealing more difficult)

#### ^C^PCR primer mix composition

2 mM of each forward and reverse primer, for all viral amplicons and 1 mM of each forward and reverse primer for the rRNA amplicon (final concentration of each primer pair in the complete 50 µL reaction was 200 and 100 nM, respectively)

#### ^D^10× PCR Top Up Buffer composition

750 mM Tris-HCl pH 8.3

200 mM (NH_4_)_2_SO_4_

1% Triton X-100

### Plate pooling

All well-barcoded PCR products from a single 96-well plate were pooled, typically 20 µL of each reaction was combined in a plastic reservoir using a multi-channel pipette, and after mixing thoroughly 1 mL was transferred to an Eppendorf tube. This was repeated for every PCR plate. 5 µL from each plate pool were re-arrayed in a new 96-well plate and treated with 2 µL of illustra ExoProStar 1-step for 30min at 37°C followed by 15 min at 80°C to remove any left-over primer.

### Second PCR (plate indexing and addition of sequencing adaptors)

A master mix with all components listed below was distributed across a 96-well plate (37.5 µL/well). To each we added 10 µL of unique dual-indexed i5/i7 primer pairs (Custom synthesized index primers with Nextflex barcodes, arrayed in 96-well plates) and 2.5 µL of ExoProStar-treated PCR1 pool. The reactions were run for 8 cycles to add sequencing adaptors with plate barcodes.

#### Master mix composition per reaction/well (volumes in µL)

**Master mix composition per reaction/well (volumes in $\mu\text{L}$ )**
|  |  |
| --- | --- |
| 10× Sequencing-ready PCR Buffer<sup>E</sup> | 5 |
| 25 mM each dNTPs | 0.5 |
| 100 mM dUTP | 0.07 |
| Hotstart Taq Polymerase | 0.5 |
| Water | 31.43 |

#### Thermocycler program

3 min at 95°C

8 cycles of: 15 sec at 95°C, 30 sec at 65°C, 30 sec at 72°C

2 min at 72°C

Cool down to 12°C

#### ^E^10x Sequencing-ready PCR Buffer composition

750 mM Tris-HCl pH 8.3

200 mM (NH_4_)_2_SO_4_

20 mM MgCl_2_

0.1% Tween 20

### Pooling and preparation for sequencing

All samples from a 96-well plate (20µl from each well) were pooled and 250µl of pooled sample was resolved on a 2% agarose gel and 220-260 bp amplicons were excised and gel purified using Qiagen gel extraction kit.

To ensure fast turnaround, the preparation of libraries for Illumina sequencing was optimized empirically. In the first four sequencing runs, standard quality control of the library, including Qubit measurement, a size analysis and qPCR, was performed. A correlation between the concentration measurement by Qubit and the qPCR was detected. In every case the molarity determined by qPCR was 10× higher than the concentration measured by Qubit. Thus, we were able to omit the size analysis and the qPCR, which are both time consuming. The library concentration was determined by three independent Qubit measurements, the obtained value in ng/µl was multiplied by 10 and used as the molarity of the sample in nanomolar. This procedure enables us to start the sequencer within 15 min after receiving the sequencing library. Final preparation of the sequencing run happens according to Illumina’s guidelines, including denaturation of the sample, neutralization and final dilution for sequencing.

### Sequencing

Depending on the sequencer type, the following concentrations were used for sequencing: 10 pM for MiSeq V2 chemistry, 15 pM for MiSeq V3 chemistry, 2.2 pM for NextSeq550 high output and 1.3 pM for NextSeq550 medium output. In every sequencing run 10% of PhiX library were spiked-in to increase complexity. To avoid contaminations with barcodes from previous sequencing runs, the sequencers were washed with bleach according to Illumina’s guidelines before every run^15^. In addition, that to avoid cross contamination of barcodes from previous runs, in practice, even if running a smaller number of samples, having 384 plate barcodes (2nd dimension) allowed us to alternate the subsets of indices used and thereby filter against any DNA remnants from previous runs that might be in the sequencer.

### Data analysis

The NGS data (fastq.gz files) were mapped in a single pass to sets of expected amplicon sequences and to sets of expected well- and plate indices using dedicated shell and awk scripts based on string-hashing that allows for 0 or 1 mismatch per amplicon and index. During method development, different parameters were tested and optimized, including single-versus paired-end sequencing, the sequencing platforms (MiSeq vs. NextSeq), and the exact positions of the indices in the primers (and thus in the reads) and the analysis was adjusted accordingly (the analysis script we make available is compatible with the final primer- and parameter set recommended for use). For redundant dual indexing, we required the correct redundant encoding of plate and well. The i5 and i7 index reads signify the plate-indices, and parts of the forward and reverse reads (in the case of paired-end sequencing) signify the well-indices. As the well-index in the forward read starts at random offsets, we first determine the amplicon identity and position, then infer the position of the well index, and finally compare the well index to the valid well index pairs; all reads with invalid plate- or well-index pairs were excluded. For the final set of primers, the offsets are made consistent for all amplicons of a given well, changing between 1 and 4 between wells such that the well-index starts between positions 2 and 5.

The analysis script is available on GitHub at https://github.com/alex-stark-imp/SARSeq and at https://starklab.org.

### Viral amplicons

The following amplicons were extracted from the NGS reads:

**Master mix composition per reaction/well (volumes in µL)**

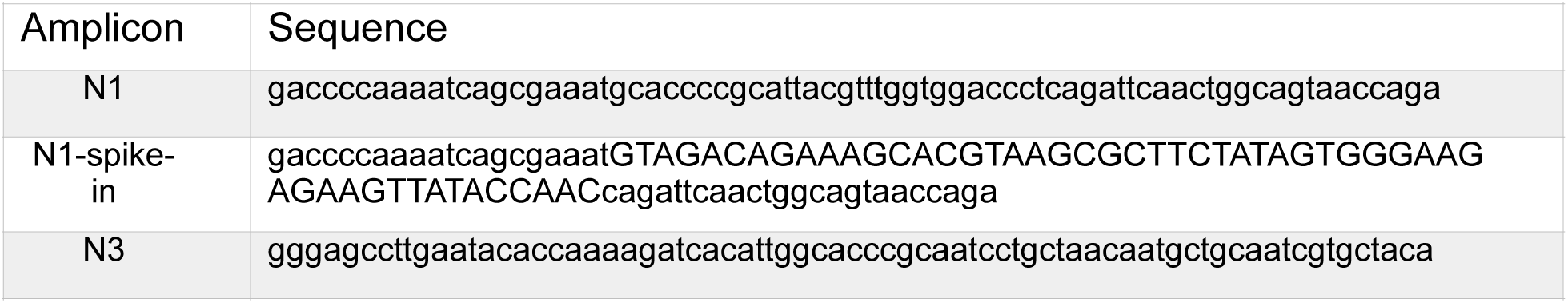
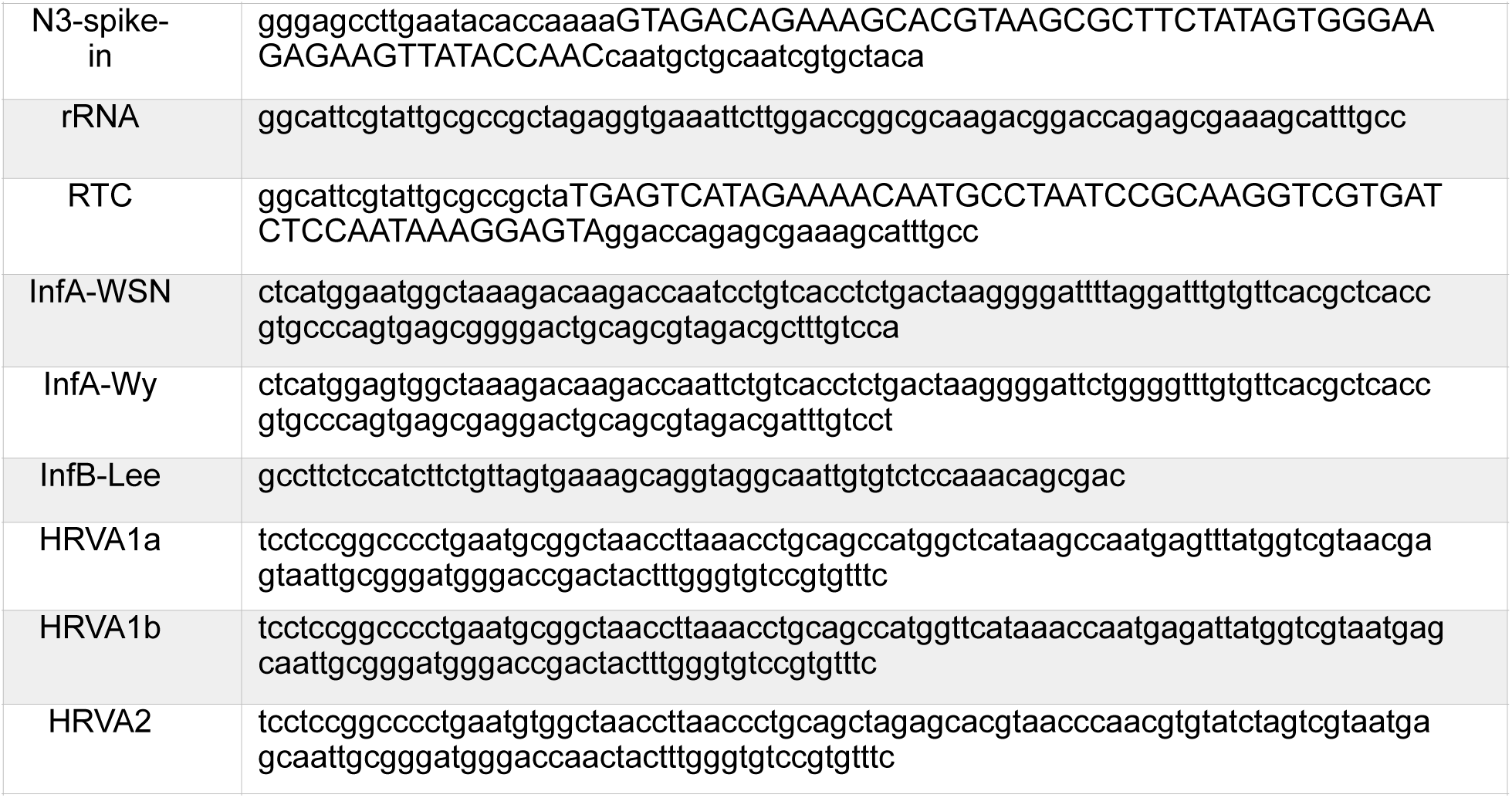

### Expression and purification of homemade enzymes

#### Reverse transcriptase 2.5

Transformed pET15b-His6-Reverse transcriptase 2.5 plasmid into E. coli strain Rosetta and plated on LB plates containing Ampicillin and Chloramphenicol. After selection, inoculated plasmid containing colony in 5ml of LB+Amp (100 g/ml) + Chloramphenicol (30 g/ml) and incubated in orbital shaker at 37°C until a visible turbidity. Transferred this inoculum to 100ml LB+ Amp + Chloramphenicol and incubated for overnight. Next day, inoculated overnight culture into 2 liters of pre-warmed LB + Amp + Chloramphenicol and incubated on orbital shaker at 37°C until OD_600_ reaches 0.5 - 0.6. Protein expression is induced by adding IPTG to 1mM final and incubated in orbital shaker at 37C for an extra 3 hours. Cells were harvested by centrifugation at 4500 rpm/15min/4°C. Resuspended bacterial pellet in 200ml of lysis buffer (40 mM Tris pH 8.0, 100 mM KCl, 10% Glycerol, 0.01% Triton X-100) supplemented with 2 X Protease Inhibitors cocktail (Roche) and 4mg/ml of lysozyme and incubated for 30 min. Sonicated suspension for 3 × 2min at 80% power of the tip. Pellet cell debris by centrifugation at 30 min at 4°C/ 20,000rpm. Transfer supernatant to a clean bottle and spin again if necessary. If there are still chunks pass supernatant through 0.45 µm filter (do not load chunky lysates on the resin). Load the clear supernatant on Ni-NTA column or beads pre-equilibrated with the lysis buffer. Wash with wash buffer. Eluted protein with the elution buffer (40 mM Tris pH 8.0, 100 mM KCl, 10% Glycerol, 0.01% Triton X-100 and 250 mM Imidazole for batch elution or 500mM for on column elution). Step elution is carried out, if column is used. Applied the Ni-NTA eluate (protein rich fraction diluted till 50mM KCl) to Mono-S column pre-equilibrated with 10 column volumes of buffer MS1 (40 mM Tris pH 8.0, 50 mM KCl,10% Glycerol, 0.01% Triton X-100, 0.1 mM EDTA and 1mM DTT). Washed the protein loaded Mono-S column with 10 column volumes of buffer MS1. Elute the reverse transcriptase with a linear gradient from 50mM to 1M KCl (buffer MS1 and buffer MS2(40 mM Tris pH 8.0, 1M KCl,10% Glycerol, 0.01% Triton X-100, 0.1 mM EDTA and 1mM DTT). Dialyzed fractions containing the protein of interest against 2 liters of dialysis buffer (40mM Tris pH 8.0, 100mM KCl, 0.1 mM EDTA, 1mM DTT, 0.01% Triton X-100 and 50% Glycerol) for overnight in the cold room. For long term storage Reverse transcriptase 2.5 is storage at −80°C and for short term at −20°C. Protein concentration was estimated with Bradford and different dilutions of the enzyme are assayed for the final optimal working concentration of reverse transcription.

#### Ribonuclease inhibitor

Ribonuclease inhibitor is expressed and purified as described previously^48^.

#### Hot-start Taq polymerase

Transformed Taq polymerase expression plasmid into E. coli strain DH5 alpha and plated on LB plates containing Ampicillin. After selection, inoculated 5 ml (LB-medium with 100 g/ml ampicillin) with a single colony of *E. coli* expressing Taq polymerase and incubated in orbital shaker at 37°C until a visible turbidity. Transferred this inoculum to 100ml LB+ Amp and incubated for overnight. Next day, inoculated overnight culture into 2 liters of pre-warmed LB + Amp and incubated on orbital shaker at 37°C until OD_600_ reaches 0.5 - 0.6. Protein expression is induced by adding IPTG to 1mM final and incubated in orbital shaker at 37C for an extra 3 hours. Cells were harvested by centrifugation at 4500 rpm/15min/4 °C and washed the cell pellet with 1X PBS. Resuspended cell pellet in 100ml Buffer A (25mM HEPES-KOH pH 7.5, 25mM Glucose, 200mM KCl, 1mM EDTA, 0.5% Tween-20 and 0.5% NP-40) with Protease inhibitors (Roche). Incubate suspension in a 250 ml Erlenmeyer flask for 1 hour at 75°C. Pellet cell debris by centrifugation at 30 min at 4°C/ 35,000 rpm and collect supernatant. Equilibrate DE-52 (DE-52, pre-swollen form, Whatman) or DEAE resin (BioRad #156-0021) by washing it 3 to 4 times with Buffer A and centrifuge 2 min at 4000 rpm, 4°C. Batch incubate the supernatant with DE-52 or DEAE for 15 min at 4°C (resin should not settle down). Centrifuge 2 min at 4000 rpm, 4°C and collected supernatant. Wash one time DE-52 or DEAE resin with 100 ml Buffer A, centrifuge for 2 min at 4000 rpm, 4°C and collect the supernatant. Both supernatant fractions combined and diluted to 40 mM KCl with Buffer B (20mM HEPES-KOH pH 7.5, 1mM EDTA, 0.5% Tween-20 and 0.5% NP-40). Apply the supernatant on a Poros 20 CM 16mmD/100mmL column (this column is not produced any longer. Therefore, if you don’t have it, you could use any strong cation exchange column). Before loading the samples equilibrate the column with 40mM KCl. After applying the sample, wash the column with 40mM KCl, till there is a stable baseline. Step elute the Taq polymerase with 300mM KCl in buffer B. Collected the peak fractions and dialyzed in dialysis buffer (20mM HEPES-KOH pH 7.5, 100mM KCl, 50% glycerol, 1mM EDTA, 0.5% Tween-20 and 0.5% NP-40 and 1mM DTT) for overnight at 4°C. Volume will reduce to about 1/3 and protein should be ready for storage (long term storage at −80°C, short term at −20°C). Measured the activity of Taq polymerase and diluted accordingly with dialysis buffer. Taq polymerase is made Hotstart compatible by using method described previously^49^.

